# Multidimensional biocircuitry of exercise adaptation: integrating *in vivo* and *ex vivo* phenomics with miRNA mapping

**DOI:** 10.1101/2025.01.20.25320845

**Authors:** Jeremy S. McAdam, Michael P. Craig, Zachary A. Graham, Brandon Peoples, S. Craig Tuggle, Regina S. Seay, Kaleen M. Lavin, Amber B. Gargus, Samia M. O’Bryan, Sufen Yang, Devin J. Drummer, Christian J. Kelley, Kalyani Peri, Margaret B. Bell, Inmaculada Aban, Gary R. Cutter, Arash Mahyari, Yuan Wen, Jin Zhang, Akshay Hira, Timothy J. Broderick, Madhavi Kadakia, Marcas M. Bamman

## Abstract

In a randomized, dose-response trial, we used molecular and phenomic profiling to compare responses to traditional (TRAD) moderate intensity endurance and resistance training vs. high-intensity tactical training (HITT) that encompassed explosive whole-body interval training and high-intensity resistance training. Ninety-four participants (18-27 years) completed 12 weeks of TRAD or HITT followed by 4 weeks of detraining. Although similar performance and body composition improvements were observed in response to HITT and TRAD, some dose-dependent differences were observed for: (i) *ex vivo* muscle tissue changes in myofiber size, capillarization, satellite cell frequency, and mitochondrial function; and (ii) differential gene expression (DGE) of muscle and serum exosomal miRNAs (miRs). However, these dose-dependent *ex vivo* muscle adaptations were overshadowed by wide-ranging inter-individual response heterogeneity (IRH). We therefore explored IRH by first establishing minimum clinically important difference (MCID) scores to classify each participant based on MCIDs for functional muscle quality (fMQ) and cardiorespiratory fitness (CRF), and then modeling all data based on MCID classification. Using higher-order singular value decomposition (HOSVD), we established multidimensional biocircuitry linked to IRH that identified the most influential features across lifestyle, body composition, performance, ex vivo muscle tissue, and miRNA mapping domains. Via cross-comparison of MCID-linked miRs identified via DGE and HOSVD, nine miRs emerged as robust features of training adaptability, providing new insights into the integrated biocircuitry driving IRH.

**NEW & NOTEWORTHY:** We examined in vivo and ex vivo adaptations to TRAD (traditional moderate-intensity endurance and resistance training) vs. HITT (high-intensity tactical training; explosive whole-body interval training and high-intensity resistance training). TRAD and HITT improved physiological performance and body composition, and induced ex vivo muscle adaptations, with remarkable inter-individual response heterogeneity (IRH) in improvements. We leveraged multidimensional modeling to identify IRH biocircuitry that integrates deep phenotyping and miR transcriptomics (serum exosomes, skeletal muscle).

## INTRODUCTION

Current public health guidelines emphasize the importance of combining endurance and resistance training to promote fitness and exercise-induced health benefits. The 2018 U.S. Department of Health and Human Services (HHS) guidelines recommend at least 150 minutes per week of moderate intensity endurance exercise or 75 minutes per week of vigorous higher intensity endurance exercise, combined with 2 or more days per week of resistance exercise targeting all major muscle groups (1). These endurance prescriptions typically focus on continuous, steady state endurance exercise.

As an alternative to continuous endurance training, high-intensity interval training (HIIT) has gained popularity for its time-efficient approach to achieving rapid fitness improvements. HIIT typically involves maximal or near maximal sprint intervals separated by brief, lower intensity recovery periods. A modified form of HIIT integrates circuits of explosive bodyweight movements and exercises with external resistance, executed with minimal rest. This approach engages more muscle groups, challenges balance and coordination, and aims to provide combined endurance and strength/power benefits. Such programs are widely used among athletes and in the military, including the US Marine Corps’ High-Intensity Tactical Training (HITT).

For endurance exercise, single-mode comparative efficacy trials of moderate-intensity continuous training (MICT) vs. HIIT are numerous and range from healthy adults to chronic disease (2–10). Studies of combined endurance and resistance training are limited and comparison to HITT is lacking. To fill this void, we conducted a randomized, comparative efficacy trial to compare TRAD and HITT over multiple weeks of training and detraining designed to deliver three major aims. Our first aim was to assess the effects of 12 weeks (3 d/wk) of traditional (TRAD) combined training versus HITT on *in vivo* physiological performance and body composition along with *ex vivo* muscle histology and mitochondrial function. These were linked to microRNA (miR) regulatory networks to map epigenetic adaptations. Training protocols were distinct stressors designed to elicit adaptations typical of both endurance and resistance training. Randomization was stratified by sex (assigned at birth) to account for sex as a biological variable. Our second aim sought to explore inter-individual response heterogeneity (IRH) by defining multidimensional biocircuits linked to IRH by integrating over 37,000 features consisting of phenomic and time series miR profiles from serum-derived exosomes and skeletal muscle in both untrained and trained states. Our third aim was to define early phase (4 wk) detraining in muscle and parallel miR expression. In a participant subset, we previously published transcriptomic responses in serum extracellular vesicles (EVs) and skeletal muscle after a single bout of TRAD or HITT in the untrained state (11).

Our identification of miRs and latent variables associated with training adaptability provide insights into the individual responses to training, potentially serving as a basis for more precise and efficient strategies for improving clinically important outcomes such as muscle quality and cardiorespiratory fitness. Further, our findings contribute to the understanding of the molecular mechanisms behind exercise adaptation that are central to athletic performance, rehabilitation, and overall health.

## MATERIALS AND METHODS

### Study Design, Participant Recruitment, Enrollment and Randomization

The Precision High-Intensity Training Through Epigenetics (PHITE) clinical efficacy trial (Department of Defense, Office of Naval Research grant: N000141613159, ClinicalTrials.gov Identifier: NCT03380923) was a parallel, randomized, two-arm, exercise dose-response efficacy trial to investigate acute molecular responses and training adaptations to two distinct prescriptions of combined exercise training in young adults. The 16-week protocol (summarized graphically in **Figure 1**) was divided into two phases: (1) a 12-week progressive exercise training phase with 3 training sessions per week followed immediately by a (2) 4-week detraining phase. *In vivo* physical performance, physiological phenotyping and *ex vivo* tissue and molecular phenotyping were performed pre- and post-training. Molecular profiling was performed using serially collected biospecimens collected in both the untrained (Week 0, pre) and trained (Week 12, post) states. *Ex vivo* tissue and molecular phenotyping was repeated following the 4-week detraining phase. The PHITE trial was approved by the University of Alabama at Birmingham (UAB) Institutional Review Board for Human Use (Protocol: F160512012) and conducted in accordance with the Declaration of Helsinki. Clinical trial registration was completed prior to recruitment of the first subject. Healthy, young adult female and male civilians aged 18-27 years were recruited from the greater Birmingham, Alabama catchment area. Participants were exercise-naïve (*i.e.*, no history of regular exercise training in the past 12 months) but could tolerate exercise without onset of angina/ischemia. Participant exclusion criteria are described in NCT03380923. All screening, enrollment/randomization, testing, exercise training, and biospecimen collections were completed at the UAB Center for Exercise Medicine (UCEM) in Birmingham, Alabama. Participants completed the US Marine Corps’ Initial Strength Test (IST), Multi-dimensional Aptitude Battery II (MABII) cognitive performance test and other baseline testing, followed by randomization to TRAD or HITT. Randomization was stratified by sex to attain sex balance and enable sex as a biological variable to serve as a valuable input feature into modeling of inter-individual response heterogeneity. A consort diagram summarizing recruitment, enrollment, and retention across the phases of the trial is presented in **Figure 2A**.

**Figure 1.**
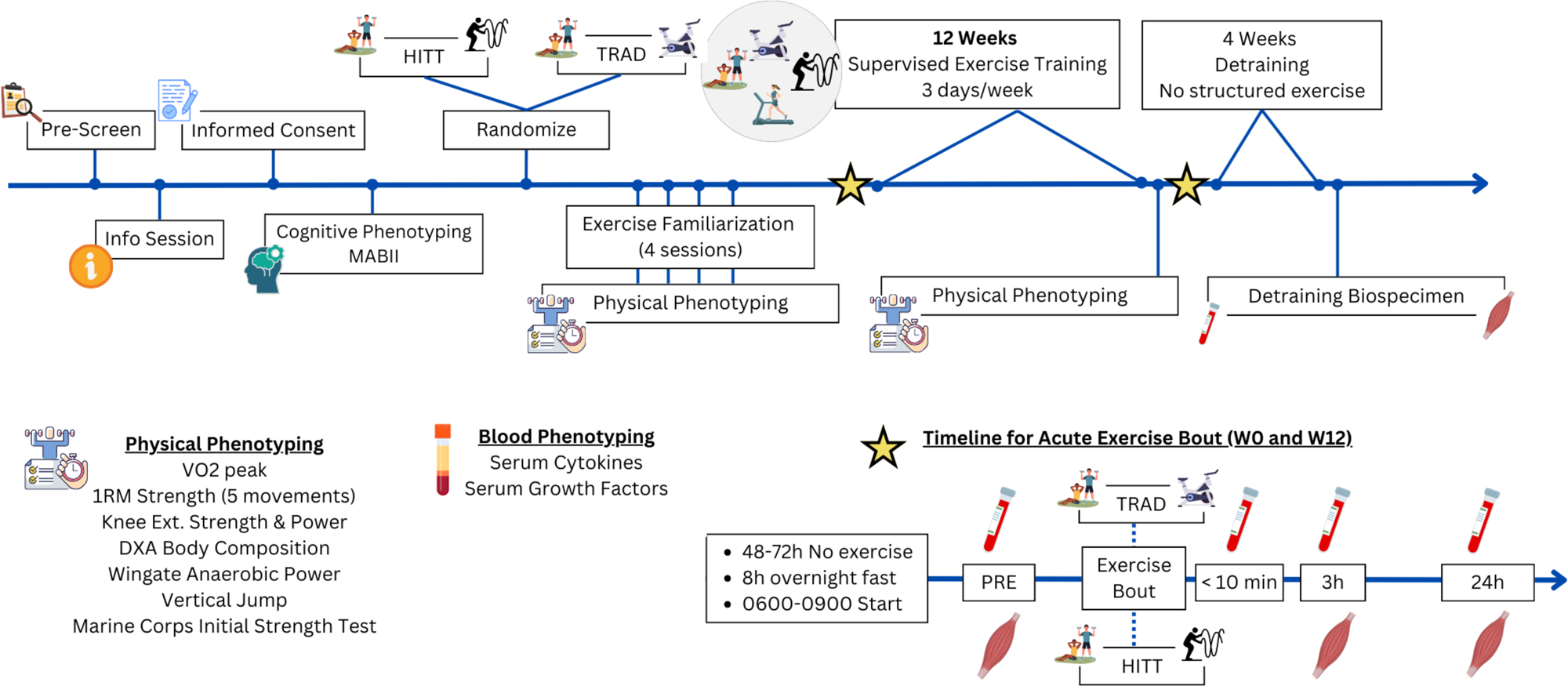
Clinical protocol for the PHITE study. TRAD: Traditional endurance plus resistance training; HITT: high-intensity tactical training. MAB II: Multidimensional Aptitude Battery-II; VO2 peak: peak oxygen uptake; DXA: Dual-energy X-ray absorptiometry; CBC: Complete blood count; 1RM: one-repetition maximum strength test.

**Figure 2.**
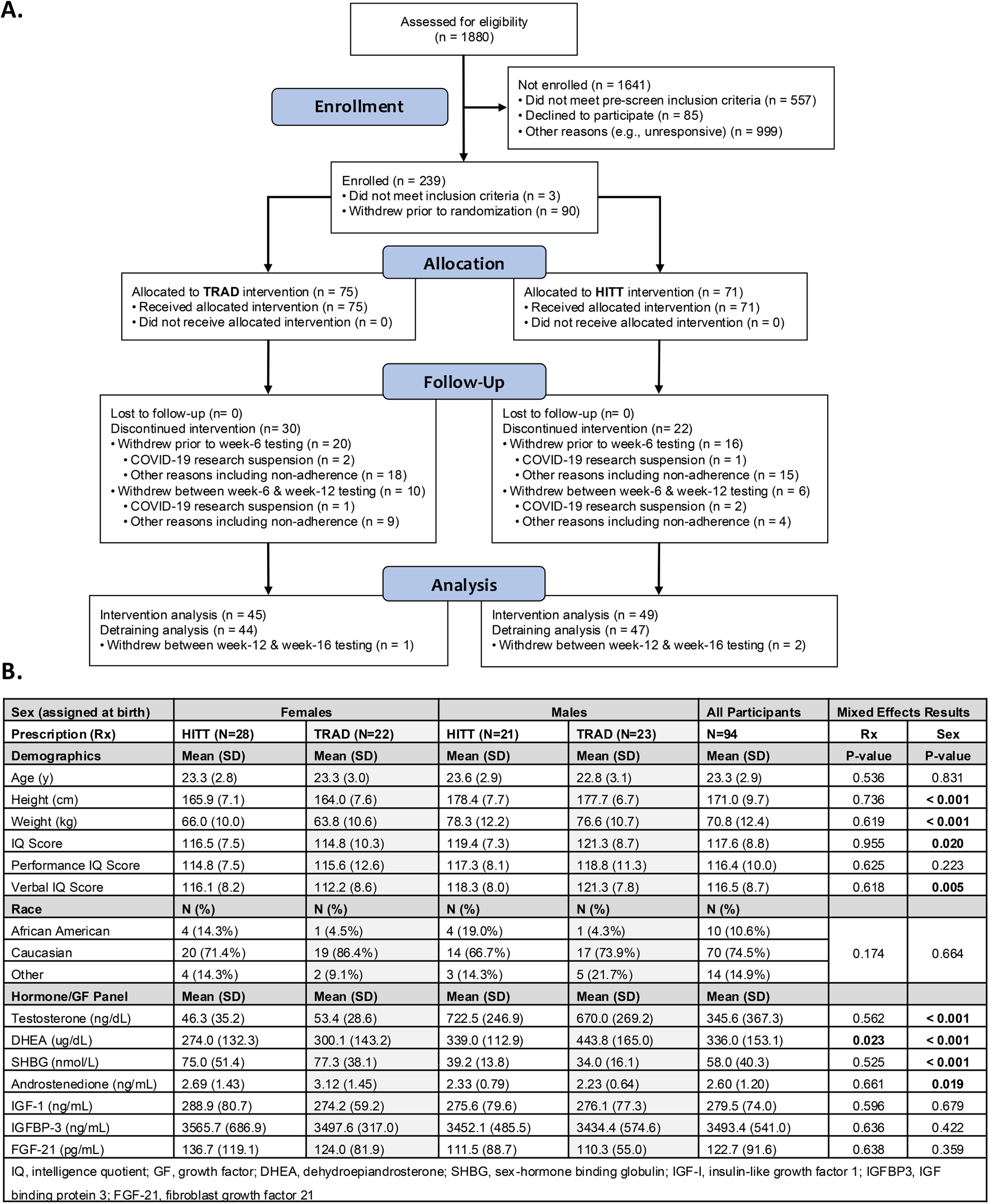
Consort diagram (A) and participant characteristics (B).

### Familiarization and Exercise Training

Prior to the first acute response exercise bout and subsequent exercise training, participants underwent four familiarization sessions to establish proper exercise technique and to progress volume and intensity to mitigate muscle soreness/damage/inflammation. The first full volume and intensity bout was the acute response testing bout described below. Participants then completed 12 weeks of supervised, progressive, combined exercise training.

### TRAD

Participants randomized to TRAD completed 30 min of steady-state, moderate intensity continuous exercise training (MICT) 3x/week (M, W, F) at a target of 70% heart rate reserve (HRR) using a LifeFitness 95CE Discover SE cycle ergometer (2x per week) or treadmill (1x per week). HRR was defined by resting HR and peak HR determined during the baseline VO_2peak_ test. Twice per week (M, F), this MICT was followed by moderate intensity whole-body resistance exercise training. Volume and intensity were prescribed as 3 sets per movement with loads targeting 13-repetition maximum (13RM) across nine movements: (chest press, seated row, overhead press, lat pulldown, triceps extension, biceps curl, calf press, squat or leg press, knee extension) plus 3 sets of abdominal crunches to volitional fatigue. Sets were separated by ∼60 sec rest. For MICT, workloads on the stationary cycle and treadmill were increased as needed to maintain 70% HRR. For resistance training, external loads were increased as needed to maintain 11-15RM, targeting 13RM. The TRAD combined exercise bout duration (M, F) was ∼90 min while the MICT-only session each W was completed in ∼40 min.

### HITT

HITT focused on explosive movements and sustained near-maximal to maximal power during 10 rounds of 30-sec exercise intervals separated by 30 seconds of rest. The 10-round circuit consisted of 9 movements 3x/week (M, W, F): rowing sprint, box jumps, burpees, split squat jumps, kettlebell swings, cycling sprints, battle ropes, wall balls, and dips. The 30-sec cycling sprint was performed twice in the 10-round circuit. Resistance training was performed 2x per week (M, F) executing the same movements as TRAD but: (i) loading intensity was higher (9RM vs. 13RM); (ii) movements were performed as “supersets” with two movements recruiting antagonistic or unaffected muscle groups completed in succession with no rest; and (iii) only 30 sec rest intervals between supersets. For the resistance training component of HITT, external loads were increased as needed to maintain 8-10RM, targeting 9RM. The HITT combined exercise bout duration (M, F) was ∼45 minutes while the HITT-only session each W was completed in ∼15 min.

### Acute Response Studies

Prior to and at the conclusion of the 12-week training program, participants completed an acute response exercise bout with serial biospecimen collections to map acute and longer-term molecular responses to TRAD vs. HITT (summarized graphically in **figure 1**). Timing for acute response studies was standardized pre- and post-training for each participant and performed in the morning fasted state (8-10 hour fast). Pre-exercise acute bout biospecimens were collected after a supine rest period of 15 minutes. The participant then completed a supervised exercise training bout of combinatorial exercise (M, F prescription) based on their randomization (HITT or TRAD). Blood draws were conducted immediately (within 10 min), 3h, and 24h post-exercise and muscle biopsies were performed 3h and 24h post-exercise. Participants consumed a nutrition shake (Ensure Plus, 64905 or Ensure 67453 for vegetarian participants) after the immediate post-exercise blood draw. Participants returned in the morning fasted state for 24h post-exercise blood and muscle collections.

### Detraining

At the conclusion of the 12-week training, participants were instructed to continue normal daily activities but to abstain from organized or structured exercise for four weeks. Participants returned for final biospecimen (muscle and blood) collection, in the morning fasted state, at the end of the four-week detraining period.

### Clinical and Performance Phenotyping

#### Multidimensional Aptitude Battery-II (MAB-II)

The MAB-II was administered using Sigma Assessment Systems (Sigma Inc., Port Huron, MI, USA) to evaluate aptitude/intelligence at baseline. MAB-II uses ten domains (information, comprehension, arithmetic, similarities, vocabulary, digit symbol, picture completion, spatial, picture arrangement, object assembly) to score verbal, performance, and full-scale IQ.

#### Body composition

Body composition was assessed via dual-energy x-ray absorptiometry (DXA) using a GE Lunar iDXA (GE Healthcare, Chicago, IL, USA, Encore v18 software) pre- and post-training. Total body scans occurred at least 24h prior to or after exposure to exercise and after a minimum 2h fast. Analyses included total body variables and partitioned, regional variables (e.g., thigh lean mass, android fat, etc)

#### Aerobic power (VO_2_peak)

VO_2_peak was determined pre- and post-training using a continuous ramp protocol on a LODE Excalibur Sport (LODE B.V., Groningen, NL) cycle ergometer. Initial resistance was set at 50 watts for females and 75 watts for males and increased 8.3 watts/min. for the first 9 minutes of the test. Thereafter the wattage increased 15 watts/min for females and 25 w/min for males until volitional fatigue or the participant was unable to maintain a cadence above 60 RPM. The main VO_2_peak achievement criteria were volitional fatigue and achieving a respiratory exchange ratio (RER) > 1.10. The minimum RER attained across all W0 and W12 VO_2_peak tests was 1.12 and the mean was 1.29. Heart rate was measured using 3 lead ECG, blood pressure was collected every other minute (even minutes) alternating with RPE (odd minutes).

#### Anaerobic power

Anaerobic power and fatigue resistance were measured using the standardized 30 s Wingate cycle test on either a LODE Excalibur Sport (LODE B.V., Groningen, NL) or Monark (Monark Exercise, Vansbro, SE) cycle ergometer pre- and post-training. Participants performed a 5 min warm-up with no load prior to the test. Per standard protocol, males were tested at a resistance of 0.7 kiloponds per kilogram bodyweight and females were tested at a resistance of 0.67 kiloponds per kilogram.

#### One-repetition maximum (1RM) strength testing

1RM was assessed for chest press, back squat, lat pulldown, knee extension, and overhead press pre- and post-training. Participants warmed up on a cycle ergometer for at least 5 min prior to 1RM testing. The order of movements for testing was standardized to reduce muscle fatigue in agonistic movements and optimize recovery between 1RM tests. For each movement participants performed 8-12 reps with a light load, 3-5 repetitions with a moderate load, 2-3 reps with a moderately heavy load, then proceeded to 1RM testing. There were 1-2 min rest intervals between warmup sets and 2 minutes between 1RM attempts. 1RM was defined as the heaviest load lifted through a full range of motion preceding two failed attempts.

#### Knee extensor isometric strength and power

Peak unilateral maximum voluntary isometric contraction (MVIC) strength and dynamic power were evaluated on the dominant leg pre- and post-training using a Biodex System 4 Pro dynamometer (Biodex Inc., Shirley, NY, USA). To determine MVIC, three 5-s maximal knee extension contractions separated by 60 sec. rest, were performed at a fixed joint angle of 60°. MVIC was defined as the peak torque generated during these contractions. Peak knee extension power was determined against a constant external resistance set at 40% of the MVIC torque. Participants completed five consecutive, full range of motion unilateral repetitions as rapidly as possible during the concentric phase (followed by a submaximal eccentric phase back to the starting position).

#### U.S. Marine Corps Initial Strength Test (IST)

The IST was administered pre- and post-training in this specific order: (i) number of successful abdominal crunches in 2 minutes; (ii) number of successful pull ups in 2 min; and (iii) time to complete a 1.5-mile run. Participants underwent a supervised treadmill warmup for 10 min. and were instructed on proper technique and evaluation criteria for abdominal crunches and pull ups. The 1.5-mile run was conducted on a Life Fitness Elevation Series Treadmill (Life Fitness, INC, Rosemont, IL, USA) at 1% grade. Participants were encouraged to complete the test as quickly as attainable and were allowed to adjust the speed as needed until the 1.5-mile distance was completed. Results from each assessment were scored by current Marine Corps IST standards.

### Biospecimen Collection and Processing

Nine serum samples per participant were collected at timepoints as indicated in Figure 1. The pre- and post-training acute response bouts each included four blood draws: (i) fasted morning resting; (ii) immediately (within 10 min) post-exercise; (iii) 3h post-exercise; (iv) 24h post-exercise. The ninth blood draw occurred after 4 wk of detraining in the fasted morning resting state. In parallel, a total of seven *vastus lateralis* skeletal muscle biopsies were performed per participant for timepoints mirroring serum collection but excluding the 10 min post-exercise timepoint as shown in Figure 1. Seven serial vastus lateralis muscle biopsies, ∼200-350mg each, were performed by alternating thighs each subsequent biopsy with spacing 1-2cm proximal to the previous biopsy site. Muscle tissue was processed and stored for immunohistochemistry or snap-frozen in ∼10-30mg aliquots for analysis of miR content and for other molecular and biochemical experiments.

### Serum Hormone and Cytokine Analyses

#### Systemic cytokine levels

To assess the potential effects of TRAD or HITT on systemic inflammation, circulating cytokine concentrations were determined in fasted, morning serum samples at baseline and post-training with the 7-plex MSD V-Plex Proinflammatory Panel I multiplex kit (MesoScale Discovery, Rockville, MD) using a MESO SECTOR S 600 according to manufacturer’s guidelines (Rockville, MD) as previously described (12). The 7-plex cytokine profile included: interferon gamma (IFN-y), interleukin (IL)-10, IL-12 p70, IL-1β, IL-6, IL-8 and tumor necrosis factor-alpha (TNF-α). All serum samples were analyzed across three plate assays with the two timepoints per participant analyzed on the same plate.

Minimum assay sensitivities for each cytokine across the three plates were: IFN-y, 0.27 – 0.42 pg/mL; IL-10, 0.025 – 0.07 pg/mL, IL-12 p70, 0.07 – 0.17 pg/mL; IL-1β, 0.028 – 0.06 pg/mL; IL-6, 0.05 – 0.20 pg/mL; IL-8, 0.03 – 0.06 pg/mL; and TNF-α, 0.12 – 0.32 pg/mL. Values below detectable limits were removed. Intra-assay and inter-assay CV% averaged, respectively: IFN-y, 3.84%, 8.62%; IL-10, 3.37%, 7.91%, IL-12 p70, 16.09%, 6.53%; IL-1β, 47.32%, 1.17%; IL-6, 7.91%, 3.92%; IL-8, 3.76%, 7.49%; and TNF-α, 3.92%, 12.09%. IL-1β was not analyzed statistically because 75% of the values were below detectable limits.

#### Anabolic hormone and growth factor profiles

As part of baseline phenotype profiling, a targeted panel of anabolic hormones and growth factors were assayed in fasted, morning serum samples. Total testosterone (total T), androstenedione, dehydroepiandrosterone sulfate (DHEA-S), and sex hormone binding globulin (SHBG) were measured on a TOSOH Bioscience AIA900 (South San Francisco, CA). Fibroblast growth factor (FGF)-21 (Millipore, Billerica, MA), insulin-like growth factor (IGF)-1 (ALPCO, Salem, NH) and IGF binding protein (BP)-3 (ALPCO) were measured using single-target ELISA kits as previously described (13). All serum samples were across two assay runs per target. Minimum assay sensitivities across the two runs averaged: total T, 10 ng/dL; DHEA-S, 4.48 µg/dL; SHBG, 0.1 nmol/L; IGF-I, 23 ng/mL; IGFBP-3, 101 ng/mL; FGF-21, 31 pg/mL; and androstenedione, 0.1 ng/mL. Intra-assay CV% averaged: total T, 6.92%; DHEA-S, 7.10%; SHBG, 6.17%; IGF-I, 5.5%; IGFBP-3, 2.93%; FGF-21, 2.68% and androstenedione, 3.84%.

### Skeletal Muscle Histology

#### General histology methods

Immunohistological protocols were performed on fasted morning biopsies collected pre-training, post-training and after detraining. 6 µm sections were prepared via a cryostat microtome (Leica, Buffalo Grove, IL) at approximately -22°C and stored at -80°C until staining. Separate sections were cut to determine: 1) myofiber type distribution and type-specific cross-sectional area (CSA); 2) capillary density, capillary contacts per type I myofiber, and capillary contacts per type 2 myofiber; and 3) number of Pax7+ satellite cells (SCs) per 100 type I and 100 type II myofibers using our well-established protocols (14–17). All time points within a given participant were immunostained and imaged together to ensure all steps and reagents including antibody concentrations, incubation times, image capture protocols, etc. were standardized.

#### Myofiber typing and sizing via myosin heavy chain (MHC) isoform immunohistochemistry

Quantitation of MHCI, MHCIIa, MHCIIx/IIax and MHCI/IIa myofibers was performed according to our established methods (18). Primary antibodies utilized were mouse anti-MHCI [0.9 µg/ml BA-D5; Developmental Studies Hybridoma Bank, University of Iowa, Iowa City, IA], mouse anti-MHCIIa (1.5 µg/ml A4.74; DSHB), and rat anti-laminin (5 μg/mL, MA1-06100; ThermoFisher). Alexa Fluor 594 goat anti-mouse (A-21145; ThermoFisher Scientific, Waltham, MA), Alexa Fluor 488 goat anti-mouse, 5 μg/mL, A-21121; ThermoFisher) and Alexa Fluor 647 goat anti-rat, 5 μg/mL, A-21247; ThermoFisher) secondary antibodies were used, sections were DNA counter-stained with DAPI-containing mounting media (VectaShield) for imaging.

#### Capillary histochemistry

Cryosections were stained using the fluorophore-conjugated ulex europaeus agglutin-1 (UEA-1 1:100, FL1061-FITC conjugate, Vector Laboratories, Burlingame, CA) to detect vascular endothelial cells. Frozen sections were stained using a MHC I and laminin cocktail and a secondary antibody cocktail containing AlexaFluor 594 (anti-MHC1), AlexaFluor 647 (anti-laminin), and UEA-I. Sections where washed and then cover-slipped with DAPI-containing mounting media (VectaShield) and imaged.

#### Pax7+ muscle stem (satellite) cell immunohistochemistry

Frozen cryosections were blocked from endogenous peroxidases with 3.0% hydrogen peroxide in PBS from the SuperBoost kit (Invitrogen) for 10 min at room temperature. Sections were stained with 1:100 Pax7+ primary antibody (Developmental Studies Hybridoma Bank, cat#: PAX7) and 1:1000 biotinylated goat anti-mouse secondary antibody (Jackson ImmunoResearch). Staining was detected using 1:100 of HRP-conjugated streptavidin in PBS from SuperBoost kit and amplified with an incubation of 1:300 Alexa Fluor 488 tyramide reagent in amplification buffer (SuperBoost). Slides were again washed and re-blocked, then incubated in laminin and MHC I antibodies as described above. After staining, sections were cover slipped, sealed, and stored at -20°C in the dark until imaged.

#### Image analysis

All images were captured with a widefield fluorescence microscope (BX51; Olympus, Tokyo, Japan) at 20x magnification with an XM10 camera (Olympus) using automated image stitching using CellSens Dimension (Olympus). Myofiber type and fCSA were assessed using MyoVision (19) with the following parameters: fiber circularity > 0.50, fCSA range of 800-25,000 µm^2^, and removal of peripheral edge fibers by moving the detected border of the entire tissue cross-section inward 30 µm. Capillary and Pax7+ satellite cell counts were performed manually by blinded laboratory staff using Image Pro 9 (Media Cybernetics, Rockville, MD) via our established protocols (20).

### Skeletal Muscle Mitochondrial Function Assays

#### Mitochondrial fractionation

Subsarcolemmal (SS) and intermyofibrillar (IMF) mitochondria were isolated using a modification of Rasmussen *et. al.* (21). Snap-frozen muscle samples from fasted morning biopsies collected pre-training, post-training and after detraining (30-50 mg) were pulverized by hand using a liquid nitrogen-cooled mortar and pestle then homogenized in a 20:1 volume-to-weight solution of ice-cold Chappell-Perry isolation buffer (100 mM KCl, 50 mM Tris-HCl, 1 mM Na-ATP, 5 mM MgSO4, 0.1 mM EGTA, 0.2% BSA, pH 7.4) + protease inhibitor cocktail (CPRoche, mini-complete; CP+PIC).

Homogenization was done at 990 rpm at 4°C using a customized Wheaton mortar and pestle and recirculating cooling system. SS were isolated by centrifuging the muscle homogenate at 600 g for 10 min at 4°C (Spin 1) then transferring the supernatant to a separate ice-cold tube for further centrifugation at 10,000 x g for 10 min at 4°C (Spin 2) to yield the SS pellet. To maximize SS mitochondrial quantity, the pellet from Spin 1 was re-homogenized with the supernatant from Spin 2, after which Spins 1 and 2 were repeated. The supernatant was removed and saved for IMF isolation and the SS-enriched pellets obtained from both rounds of homogenization were resuspended with 30-50 µl CP+PIC and used immediately or stored at -80°C. The remaining supernatant was combined with the pellet from Spin 1 and a protease (*Bacillus* sp. Type XXVII, Sigma-Aldrich) was added to release the IMF fraction. Samples were re-homogenized then centrifuged using the parameters of Spin 2. The IMF mitochondrial enriched pellet was resuspended with 30-50 µl CP+PIC and used immediately or stored at -80°C.

#### Mitochondrial enzyme activity assays

SS and IMF mitochondrial enzyme activities were measured as we have described (22). In brief, prior to assessment of mitochondrial complex activities, protein concentration was determined using a Lowry assay in duplicate. Complex I activity was immediately measured on a DU800 spectrophotometer using 2,6-dichloroindophenol (DCIP) at 600 nm. Complex IV activity was measured by the oxidation of cytochrome c at 550 nm. Complex I and IV data are represented as the pseudo first order rate constant (*k*) divided by protein concentration. Citrate synthase (CS) was measured as a surrogate index of mitochondrial volume using a coupled reaction with oxaloacetate, acetyl-CoA, and 5,5-dithiobis-(2,4-nitrobenzoic acid). Complex I and IV activities were adjusted for total CS in that given fraction (SS or IMF).

### Muscle Total RNA and Small RNA Enrichment

Total RNA was isolated from ∼25mg of snap frozen muscle tissue using the mirVana^TM^ Paris Kit (Thermo Fisher Scientific, Carlsbad, CA, catalog # AM1556), treated with DNase and checked for genomic DNA contamination by qPCR for GAPDH. RNA was quantified by Small RNA Agilent analysis and reverse transcribed to cDNA using adapter-specific Ion RT primers v2 (Life Technologies, Carlsbad, CA), and enriched for small RNA using two rounds of size selection ethanol washes.

### Serum Exosome Isolation and miR Extraction

Exosomes were isolated from 1 mL of serum thawed to room temperature using the Total Exosome Isolation (from serum) kit (Life Technologies), followed by exosomal RNA isolation using the Total Exosome RNA and Protein Isolation Kit (Life Technologies, catalog # 4478545). Exosomal RNA was then converted to cDNA directly using adapter-specific Ion RT primers v2 (Life Technologies, Carlsbad, CA) without Agilent quantitation or enrichment for small RNA.

### Small RNA Sequencing

Small RNA-Seq was performed and as described previously (23) using the Ion-Torrent platform to identify microRNAs from peripheral which are differentially expressed in serum exosomes and skeletal muscle in response to exercise. Briefly, cDNA samples from both muscle and exosomes were barcoded using Platinum PCR SuperMix High Fidelity polymerase with IonXpress RNA 3’ Barcode primer and unique 5’ Ion Xpress RNA-Seq Barcode Primers using the Ion Xpress RNA-Seq Barcode 1-16 Kit (Life Technologies, Carlsbad, CA). The yield and size distribution of the cDNA libraries were assessed using the Agilent 2100 Bioanalyzer DNA1000 chip (Agilent Technologies, Santa Clara, CA). Total barcoded cDNA within the 50 – 300 base pair range was considered to be derived from small RNA. 7.5 picomoles each of 8-10 barcoded libraries were pooled and clonally amplified onto Ion Sphere^TM^ Particles (ISPs) according to the manufacturer’s protocol (Ion PI Template OneTouch^TM^ 200 Template Kit v3) and enriched using the Ion OneTouch 2 ES system (Life Technologies, Carlsbad, CA). Small RNA-Seq was performed using the Ion-Torrent platform to identify microRNAs from peripheral which are differentially expressed in serum exosomes and skeletal muscle in response to exercise. Enriched ISPs were sequenced on the Ion Proton Next Generation Sequencing system using the Ion P1 chip v2 Kit and Ion PI^TM^ Sequencing 200 kit v3 (Life Technologies, Carlsbad, CA) with 500 sequencing flows.

### miR Differential Expression Analyses

RNA-Seq data processing was performed using Partek Flow v10.0.21.0706 (Partek Inc., Chesterfield, MO). Samples with total read counts less than 200,000 were re-sequenced and the resulting FASTQ files were merged in Partek Flow. Data analysis was performed as described in Sakaram et. al. (23). Briefly, unaligned reads were trimmed to a fixed length based on an average PHRED quality score greater than 20, aligned to miRBase mature miRs version 21 of the hg38 (GRCh38) assembly using a Bowtie aligner, and quantitated using the miRBase version 21 reference. Raw read counts were normalized using the Trimmed Mean of M values (TMM) normalization method with an offset of 1.0 to enable downstream differential expression analysis. TMM normalized read counts and associated raw FASTQ files are available at GSE269533.

### Differential Gene Expression (DGE) Analysis

Differential gene expression analysis was performed to identify exercise signatures of the exercise and detraining responses in the full unsegmented dataset and after segmentation on sex (*i.e.,* male and female), exercise cohort (*i.e.*, TRAD and HITT) and by individual response (i.e. fMQ+/CRF+ vs all non-fMQ+/CRF+. MiR showing differential expression in the untrained (W0) and trained (W12) states were identified by comparing the 3hr “acute” and 24hr timepoints to the corresponding “pre” baseline sample. Training-specific effects were identified by comparing the baseline trained (W12 pre) samples to untrained baseline (W0 pre) samples. Detraining effects were identified by comparing the detrained W16 samples to the trained W12 pre baseline samples. Fold changes and unadjusted p-values were determined using the Lognormal with Shrinkage (LNS) model for Gene Specific Analysis (GSA) in Partek Flow software, v10.0 (2020). Significant differences in miR levels were defined as absolute fold-changes ≥ 1.5, p ≤ 0.05 and average read count values ≥ 10 in the categories being compared. Detailed fold-change, p-value and average read count information for all DE miR is provided in **Table S1**. UpSet plots were generated in GraphPad Prism version 10.2.1 for Windows (GraphPad Software, Boston, Massachusetts USA, www.graphpad.com).

### miR Target Analysis

mRNA targets of microRNAs showing differential expression were identified using MirTarbase Release 9.0 (24). mRNA targets with weak mean-target interaction (MTI) evidence were removed leaving only those with western blot, reporter assay or qPCR support (i.e., strong evidence). Signaling networks were generated using Cytoscape v3.10.2 (25). Core signaling networks were generated by filtering to include only mRNA targeted by 2 or more miR in the dataset.

### Pathways Analysis

Pathways analyses of mRNA targets were performed using the KEGG 2021 database via ENRICHR (26) in R with a particular focus on exercise-related pathways and excluding chronic and infectious disease pathways.

## QUANTIFICATION AND STATISTICAL ANALYSIS

### Data Management

Physical and biological phenotyping data were processed and analyzed using our four-phase data management pipeline (**Figure S1**). All data were processed and organized using R (details provided in supplemental table 1). Values were flagged as outliers and removed when they were above or below 3.6 standard deviations from the mean within each level of the fixed factors (time point). A model for each variable of interest using W0, W12 and W16 values (where applicable) was created, and the mahalanobis distances for each participant from the center of the model for that variable were calculated. The model center was determined using the minimum covariance determinant method. This method is more reliable when outliers are present. We then calculated p-values for the mahalanobis distances from a chi-square distribution and potential outliers were flagged for review when p < 0.001.

### Statistical analyses

Principal outcomes were assessed using repeated measures ANOVAs with time as the repeated measures factor and intervention (HITT vs. TRAD) as the between groups factor. An *a priori* alpha level of 0.05 was used to determine the main effects of intervention group and time, and group x time interaction. Normality of residuals was assessed using Shapiro-Wilks (W: Wilk’s Statistic) and Kolmogorov Smirnov tests. Residuals were visualized using QQ and residual plots. When normality was violated (i.e. 75% or more of the levels of the data were statistically significant), data were square root transformed, and normality was re-tested. Homogeneity of variance between groups was assessed using a Levene’s test and equality of variance across time was assessed using Maulchy’s test of sphericity. If sphericity was violated a Greenhouse-Geisser correction was used to adjust the degrees of freedom for determining the critical value for F to account for the increase in Type I error rate. For targeted muscle tissue analyses where data were collected at three timepoints (pre-training W0, post-training W12, detraining W16), we used mixed effects models. Variance was estimated using Restricted Maximum Likelihood (REML) and group and time were fixed effects. Intercept was allowed to vary across individual participants (random effect). We calculated degrees of freedom using the Satterwaite method to determine F and p values to test for differences between fixed factors. Significance for all tests was accepted at p < 0.05. Where significant interactions or main effects were found, we conducted post-hoc pairwise t-tests on planned comparisons. Where a significant main effect of time existed, we compared W0 vs. W12 and W12 vs. W16. Where significant group by time interactions were detected, we conducted between groups comparisons at W0, W12, W16 and then within groups comparisons (W0 vs. W12, W12 vs. W16) and reported unadjusted p values.

### Minimum Clinically Important Difference (MCID) Determination

Thresholds for minimally clinically important difference (MCID) scores were calculated using a distribution-based method for functional muscle quality (fMQ; one repetition maximum of bilateral knee extension normalized to bilateral thigh lean mass) and cardiorespiratory fitness (CRF; VO_2peak_ in L/min). The MCID was determined by multiplying the standard deviation of the individual percent change scores by 0.5 according to established methods (27). Individuals at or above the MCID threshold for either metric were classified as responders (+) for fMQ and/or CRF. We then created a binary classification as dual responders (fMQ+/CRF+) or non-dual responders (fMQ+/CRF- or fMQ-/CRF+ or fMQ-/CRF-). All responder analyses (differential gene expression (DGE) analysis and HOSVD) were conducted on this binary dual responder outcome (fMQ+/CRF+ vs. all others).

### Higher-Order Singular Value Decomposition (HOSVD)

To evaluate potentially important features that contribute to MCID status, we utilized Tucker decomposition (also known as Higher Order Singular Value Decomposition: HOSVD) (28) to break down high-dimensional data into interpretable components called latent variables (LVs). Phenotypic and miR data from W0 and W12 were used as input features to HOSVD as summarized in **Figure S2**. Phenomic variables that had > 20% missing values excluded from HOSVD. Missing values were imputed using the field standard MICE method (Multiple Imputation by Chained Equations) (29) using m=5 imputations and 10 iterations. Lasso regression was used primarily, but stochastic regression was also used when appropriate. Baseline miRs at W0 and W12 were included in the model as raw values, whereas miRs at 0h, 3h and 24h were included as change scores from the correspinding W0 or W12 baseline.. HOSVD decomposed the high dimensional data into three key LV matrices: time, participant, and feature LVs. We downselected to the 10 highest ranking LVs in the fMQ+/CRF+ vs. all others MCID classification. We then reduced dimensionality in two steps: 1) combined the top 5% of features from each of these top 10 LVs and eliminated any redundancies; and 2) statistically tested each feature between fMQ+/CRF+ and all others using t-tests for continuous data and Chi-Square tests for categorical input features. Welch’s t-tests were used when there was unequal variance; otherwise, an equal variance t-test was used. DiVenn networks were generated in DiVenn 1.2 (30). Canonical pathways impacted were identified using ENRICHR analysis in R as detailed in “Pathway Analysis” above using the Kyoto Encyclopedia of Genes and Genomes (KEGG 2021) pathway database (26, 31). The canonical pathway analysis was conducted on the down-selected miRs that have strong evidence in MirTarbase Release 9.0 (24), matching the approach taken in our DGE analysis.

## RESULTS

### Participant characteristics were similar across randomized prescriptions

As shown in **Figure 2A**, 94 participants completed the efficacy trial [TRAD: n=45 (22F/23M) and HITT: n=49 (28F/21M)] and 91 participants completed the detraining phase (TRAD: n=44; HITT: n=47). Baseline demographics and a targeted panel of serum anabolic/androgenic hormones and growth factors are summarized in **Figure 2B**. As expected, sex differences were observed in testosterone, DHEA, SHBG and androstenedione. Sex-stratified randomization resulted in no baseline differences between TRAD and HITT in any measurement except DHEA (p=0.023).

### HITT and TRAD prescriptions were met

Session adherence rates for the 12 weeks of 3x/wk training were high with an overall adherence of 94% ± 5% attendance (TRAD: 93.7% ± 5.3%; HITT: 94.3% ± 5.7%). The 70% HRR target intensity for the MICT component of TRAD across the 12 wk of training was achieved, averaging 70.2% ± 5.1% HRR. There was no comparable heart rate-defined intensity target for the HITT intervals, as intensity was maximum and not steady state. For the progressive RT component of each program, prescribed loading intensity was 8-10 repetitions maximum (RM) per set for HITT, and lower at 11-15 RM for TRAD. These intensities were met as exemplified by representative lower and upper body movements: squat (HITT 9.8 ± 2.1; TRAD 12.9 ± 2.3 rep/set) and chest press (HITT 9.1 ± 2.4; TRAD 11.8 ± 3.0 rep/set). Under the direct supervision of the interventionist, participants also adhered to the differentially prescribed between-set rest intervals in HITT vs TRAD, whereby HITT were given only 30 s between supersets (2 sequential movements with no rest) vs. protracted rest periods of 60 s between individual sets for TRAD (with no supersets).

### HITT and TRAD similarly improved *in vivo* physiological performance and body composition

Both TRAD and HITT yielded similar training-induced benefits in indices of health and performance. There were overall improvements in (main time effects p < 0.001) peak aerobic power (VO_2_peak, 11%), mechanical power at VO_2_peak (14%), peak anaerobic power (16%), upper and lower body strength (14-27%), and lower body power (29%) (**Table 1**). A group*time interaction (p=0.009) during the Wingate anaerobic power test indicated superior fatigue resistance in TRAD. Additionally, trends toward group*time interaction (p = 0.068) suggested greater gains in peak anaerobic power and upper body strength in HITT (chest press p = 0.063; lat pulldown p = 0.090). Improvements in body composition were comparable between TRAD and HITT, with both reducing total body fat, abdominal subcutaneous fat and visceral adipose tissue concomitant with gains in total body, thigh and arm lean mass (**Table 1**). Neither prescription induced detectable systemic inflammation (**Table 2**). Overall, TRAD and HITT were similarly efficacious in enhancing physiological performance and body composition.

**Table 1.** In vivo physiological and performance phenotyping adaptations to HITT and TRAD exercise training.

| Prescription (Rx) | HITT |  | TRAD |  | Model Results |  |  |
| --- | --- | --- | --- | --- | --- | --- | --- |
| Variable | Pre-training Mean (SD) | Post-training Mean (SD) | Pre-training Mean (SD) | Post-training Mean (SD) | Time (P-value) | Rx (P-value) | Interaction (P-value) |
| **Aerobic Power (cardiorespiratory fitness, CRF)** |  |  |  |  |  |  |  |
| VO<sub>2</sub>peak (L/min) | 2.36 (0.71) | 2.62 (0.70) | 2.35 (0.63) | 2.60 (0.68) | < **0.001** | 0.911 | 0.948 |
| VO<sub>2</sub>peak (mL/kg/min) | 33.16 (7.10) | 36.58 (7.08) | 33.16 (6.77) | 36.75 (7.04) | < **0.001** | 0.951 | 0.862 |
| Peak heart rate (bt/min) | 188.6 (9.8) | 185.1 (8.4) | 191.5 (9.7) | 186.1 (10.6) | < **0.001** | 0.376 | 0.237 |
| Peak RER (VCO<sub>2</sub>/VO<sub>2</sub>) | 1.295 (0.083) | 1.253 (0.069) | 1.288 (0.078) | 1.265 (0.059) | < **0.001** | 0.864 | 0.227 |
| Peak oxygen pulse (ml O<sub>2</sub>/beat) | 13.12 (4.94) | 14.35 (4.10) | 12.40 (3.65) | 14.73 (4.44) | < **0.001** | 0.865 | 0.144 |
| Peak power (W) | 181.1 (60.3) | 206.0 (58.6) | 184.9 (60.4) | 212.3 (63.0) | < **0.001** | 0.698 | 0.629 |
| **Anaerobic Power (30-s Wingate)** |  |  |  |  |  |  |  |
| Peak power (W) | 639.6 (198.8) | 771.9 (217.1) | 664.4 (252.5) | 741.5 (198.4) | < **0.001** | 0.949 | <u>0.068</u> |
| Peak power (W/kg) | 8.87 (2.03) | 10.60 (2.11) | 9.27 (2.61) | 10.44 (1.94) | < **0.001** | 0.779 | 0.181 |
| Rate of fatigue (%) | 54.1 (13.9) | 61.1 (12.8) | 58.1 (18.5) | 55.0 (11.5) | 0.298 | 0.661 | **0.009** |
| **Neuromuscular Strength and Power** |  |  |  |  |  |  |  |
| 1RM squat (kg) | 71.07 (29.77) | 91.50 (29.46) | 72.24 (31.50) | 89.92 (33.02) | < **0.001** | 0.975 | 0.287 |
| 1RM knee extension (kg) | 86.13 (34.91) | 107.51 (38.21) | 81.33 (28.94) | 101.72 (37.78) | < **0.001** | 0.463 | 0.792 |
| 1RM chest press (kg) | 51.96 (27.05) | 62.64 (27.65) | 50.45 (26.28) | 57.97 (26.70) | < **0.001** | 0.582 | <u>0.063</u> |
| 1RM overhead press (kg) | 36.87 (15.72) | 41.46 (16.59) | 35.37 (16.48) | 40.99 (16.36) | < **0.001** | 0.770 | 0.432 |
| 1RM lat pulldown (kg) | 51.21 (17.56) | 59.39 (18.61) | 47.44 (20.31) | 53.52 (19.71) | < **0.001** | 0.225 | <u>0.090</u> |
| Unilateral knee extension MVIC (N·m) | 196.4 (70.0) | 221.4 (75.2) | 183.9 (57.4) | 210.2 (65.9) | < **0.001** | 0.393 | 0.842 |
| Unilateral knee extension power (W) | 183.0 (96.9) | 228.3 (110.7) | 157.5 (92.0) | 211.0 (97.6) | < **0.001** | 0.278 | 0.590 |
| **Body Composition** |  |  |  |  |  |  |  |
| Total lean mass (kg) | 46.46 (9.75) | 47.69 (9.75) | 45.04 (9.93) | 46.35 (10.18) | < **0.001** | 0.533 | 0.796 |
| Bilateral thigh lean mass (g) | 11994 (2892) | 12330 (2664) | 11503 (2749) | 11953 (2827) | < **0.001** | 0.465 | 0.514 |
| Bilateral arm lean mass (g) | 5432 (1805) | 5658 (1829) | 5379 (1896) | 5582 (1922) | < **0.001** | 0.874 | 0.685 |
| Total fat mass (kg) | 21.68 (7.31) | 20.64 (6.90) | 21.18 (7.09) | 20.23 (7.36) | < **0.001** | 0.775 | 0.823 |
| Visceral fat mass (g) | 333 (288) | 295 (284) | 307 (266) | 295 (253) | **0.004** | 0.823 | 0.129 |
| Subcutaneous abdominal fat mass (g) | 1179 (690) | 1102 (667) | 1255 (623) | 1133 (636) | < **0.001** | 0.704 | 0.215 |
| Bodyfat percentage (%) | 30.50 (8.51) | 29.03 (8.26) | 30.75 (8.58) | 29.18 (8.99) | < **0.001** | 0.915 | 0.773 |

HITT: High-Intensity Tactical Training consisting of explosive whole-body interval training and high-intensity resistance training; TRAD: Traditional moderate intensity endurance and resistance training. Rx: exercise prescription. Time: main effect of time (week 0 vs. after 12 weeks of exercise); Interaction: Rx by Time interaction p value; SD: standard deviation. VO<sub>2</sub>peak: peak oxygen consumption; RER: respiratory exchange ratio; 1RM: one repetition maximum; MVIC: maximal voluntary isometric contraction; bt/min: beats per minute; W: watts; N·m: newton meters.

**Table 2.** *Ex vivo* skeletal muscle phenotyping of adaptations to HITT and TRAD exercise training.

| Prescription (Rx) | HITT |  |  | TRAD |  |  | Mixed Effects Results |  |  |
| --- | --- | --- | --- | --- | --- | --- | --- | --- | --- |
| Variable<br>(post hoc contrasts noted) | Pre-training<br>Mean (SD) | Post-training<br>Mean (SD) | Detraining<br>Mean (SD) | Pre-training<br>Mean (SD) | Post-training<br>Mean (SD) | Detraining<br>Mean (SD) | Time<br>(P-value) | Rx<br>(P-value) | Interaction<br>(P-value) |
| **Myofiber Cross-sectional Area (fCSA, μm²)** |  |  |  |  |  |  |  |  |  |
| Type I (*†) | 3610 (950) | 3827 (1116) | 3561 (1011) | 3258 (793) | 3851 (904) | 3539 (890) | < **0.001** | 0.477 | 0.062 |
| Type II | 3815 (977) | 4263 (1224)* | 3653 (1092)† | 3471 (937) | 4162 (1059)* | 3964 (1126) | < 0.001 | 0.700 | **0.018** |
| Mean | 3825 (930) | 4111 (1048) | 3633 (990)† | 3394 (728) | 4060 (935)* | 3842 (973) | < 0.001 | 0.567 | **0.008** |
| **Myofiber Type Distribution (%)** |  |  |  |  |  |  |  |  |  |
| Type I | 31.8 (7.1) | 35.2 (8.1) | 36.0 (10.7) | 34.0 (6.7) | 33.6 (12.0) | 31.3 (9.3) | 0.108 | 0.320 | 0.083 |
| Type IIa | 62.2 (8.1) | 59.9 (8.9) | 59.2 (11.5) | 60.5 (6.8) | 61.2 (12.2) | 63.7 (9.2) | 0.546 | 0.324 | 0.179 |
| Type IIx/IIax | 2.1 (3.1) | 1.8 (2.6) | 1.8 (3.1) | 2.3 (2.9) | 2.2 (3.0) | 1.5 (2.1) | 0.107 | 0.449 | 0.562 |
| Type I/IIa | 4.0 (3.2) | 3.1 (3.3) | 3.1 (2.5) | 3.2 (2.5) | 3.0 (2.5) | 3.5 (3.7) | 0.575 | 0.649 | 0.429 |
| **Capillary** |  |  |  |  |  |  |  |  |  |
| Caps / Type I myofiber (*) | 4.19 (1.18) | 4.54 (0.99) | 4.66 (1.02) | 3.83 (0.74) | 4.59 (0.91) | 4.50 (0.79) | < **0.001** | 0.352 | 0.109 |
| Caps / Type II myofiber | 4.06 (1.20) | 4.46 (0.81)* | 4.35 (1.04) | 3.49 (0.95) | 4.44 (0.91)* | 4.45 (1.02) | < 0.001 | 0.396 | **0.018** |
| Cap density (caps/mm²) | 276.2 (96.7) | 284.4 (80.4) | 294.8 (89.26) | 268.2 (69.4) | 276.1 (72.1) | 301.1 (81.0) | 0.193 | 0.803 | 0.842 |
| **Pax7<sup>+</sup> Satellite Cells (SCs)** |  |  |  |  |  |  |  |  |  |
| SCs / 100 Type I | 2.43 (2.49) | 2.35 (1.63) | 3.30 (2.96) | 3.35 (2.36) | 5.00 (5.03)* | 2.91 (2.80)† | 0.348 | 0.022 | **0.025** |
| SCs / 100 Type II (*†) | 1.19 (0.92) | 2.03 (1.46) | 1.44 (0.875) | 1.92 (1.65) | 3.59 (2.89) | 1.73 (1.85) | < **0.001** | **0.002** | 0.124 |
| SCs / 100 myofibers | 1.69 (1.51) | 2.15 (1.32) | 1.98 (1.202) | 2.24 (1.47) | 4.33 (3.68)* | 1.99 (1.69)† | < 0.001 | 0.007 | **0.006** |
| **Subsarcolemmal (SS) and Intermyo-fibrillar (IMF) Mitochondrial Enzyme Activities** |  |  |  |  |  |  |  |  |  |
| SS CS (mU/mg) (*†) | 116.2 (74.0) | 153.0 (95.0) | 113.2 (76.8) | 111.1 (89.0) | 127.6 (92.6) | 107.1 (77.7) | **0.002** | 0.525 | 0.359 |
| SS cx I (μM/min/mU CS) | 0.223 (0.127) | 0.202 (0.131) | 0.243 (0.150) | 0.282 (0.232) | 0.348 (0.276) | 0.320 (0.259) | 0.493 | **0.011** | 0.203 |
| SS cx IV (k/sec/mU CS) | 34.60 (27.59) | 36.15 (23.15) | 33.53 (22.99) | 39.86 (34.92) | 49.08 (43.90) | 34.48 (21.45) | 0.060 | 0.240 | 0.265 |
| IMF CS (mU/mg) | 61.7 (42.2) | 89.0 (54.0)* | 63.5 (48.4)† | 62.0 (55.8) | 69.9 (61.2) | 72.9 (65.4) | 0.002 | 0.734 | **0.024** |
| IMF cx I (μM/min/mU CS) | 0.172 (0.120) | 0.190 (0.125) | 0.176 (0.126) | 0.222 (0.176) | 0.228 (0.167) | 0.251 (0.188) | 0.501 | 0.066 | 0.661 |
| IMF cx IV (k/sec/mU CS) | 15.10 (10.72) | 15.48 (12.16) | 15.35 (11.41) | 16.81 (12.96) | 16.27 (10.81) | 17.64 (10.89) | 0.827 | 0.418 | 0.985 |
| Total CS (mU/mg) (*†) | 81.6 (60.5) | 100.7 (64.4) | 85.7 (61.0) | 85.6 (69.3) | 101.8 (67.5) | 90.7 (74.0) | < **0.001** | 0.798 | 0.953 |
| **Serum Cytokines** |  |  |  |  |  |  |  |  |  |
| IFN-γ (pg/mL) | 5.042 (2.223) | 5.667 (2.560) | N/A | 6.152 (4.731) | 6.882 (5.683) | N/A | 0.234 | 0.089 | 0.926 |
| IL-10 (pg/mL) | 0.361 (0.157) | 0.418 (0.224) | N/A | 0.377 (0.270) | 0.546 (0.908) | N/A | 0.132 | 0.344 | 0.455 |
| IL-12 p70 (pg/mL) | 0.306 (0.203) | 0.328 (0.222) | N/A | 0.221 (0.102) | 0.257 (0.155) | N/A | 0.149 | 0.080 | 0.726 |
| IL-6 (pg/mL) | 0.683 (0.388) | 0.658 (0.364) | N/A | 0.662 (0.409) | 0.663 (0.443) | N/A | 0.770 | 0.922 | 0.752 |
| IL-8 (pg/mL) | 9.734 (3.070) | 10.587 (4.079) | N/A | 8.570 (2.684) | 9.049 (2.456) | N/A | 0.066 | **0.023** | 0.604 |
| TNF-α (pg/mL) | 2.644 (0.644) | 2.689 (0.662) | N/A | 2.557 (0.606) | 2.637 (0.680) | N/A | 0.194 | 0.594 | 0.705 |

HITT: High-Intensity Tactical Training consisting of explosive whole-body interval training and high-intensity resistance training; TRAD: Traditional moderate intensity endurance and resistance training. Time: main effect of time (week 0 vs. after 12 weeks of exercise); Interaction: Rx by Time interaction p value; SD: standard deviation. Cap, capillary; SCs, satellite cells; CS, citrate synthase; SS, subsarcolemmal mitochondria; IMF, intermyofibrillar mitochondria; cx, complex, IFN-γ, Interferon gamma; IL, Interleukin, TNF-α Tumor Necrosis Factor alpha. For post-hoc contrasts, symbols on the variable name localize time effects detected in the full cohort, whereas the same symbols on time points within HITT or TRAD localize the same time effects within group: \*change pre-training to post-training, p < 0.05; †change post-training to detraining, p < 0.05.

### HITT and TRAD elicited differential *ex vivo* adaptations in skeletal muscle

We observed significant group*time interactions indicating 2-fold greater myofiber hypertrophy in TRAD. Mean fiber cross-sectional area (fCSA) increased 20% in TRAD vs. 7% in HITT (p=0.008) while type II fCSA increased 20% in TRAD vs. 12% in HITT (p=0.018) (**Table 2**). A main time effect was observed for type I fCSA hypertrophy (12%) with no difference between groups (**Table 2**). Myofiber type distribution remained unchanged (**Table 2**). Muscle capillary angiogenesis increased overall, with a 14% rise in capillary contacts per type I myofiber and no differences between groups (**Table 2**). However, capillary contacts per type II myofiber increased 27% in TRAD vs. 10% in HITT. Capillary density remained unchanged in both groups, indicating capillary angiogenesis kept pace with hypertrophy. Pax7^+^ muscle satellite cells (SCs) per 100 myofibers showed a significant group*time interaction, with a 93% increase pre- to post-training in TRAD vs. 27% in HITT (p=0.006) (**Table 2**). SC frequency per 100 type I myofibers rose 49% in TRAD with no change in HITT (p=0.025), while type II-associated SCs increased similarly in both groups (80% overall). For mitochondrial function, HITT increased intermyofibrillar (IMF) citrate synthase (CS) 44% vs. 13% in TRAD (group*time, p=0.024). Subsarcolemmal (SS) CS and total CS increased similarly in both groups (**Table 2**). Normalizing complex I and IV activities to CS showed no changes, suggesting the primary effect was training-induced mitochondrial biogenesis rather than enhanced mitochondrial function. Taken together, this suggests the equal improvements in VO_2_peak were linked to distinct peripheral muscle adaptations: TRAD increased perfusion capacity through greater capillary angiogenesis, while HITT enhanced IMF mitochondrial biogenesis.

### Training state (untrained, trained) and exercise prescription (HITT, TRAD) influenced miR signatures of the acute exercise response *Muscle*

Differential gene expression (DGE) analysis compared the miR profiles of TRAD (n=43) and HITT (n=47). Pre-exercise baseline muscle samples yielded only 3 differentially expressed (DE) miRs (miR-143-3p, miR-145-5p and miR-2776-3p), supporting sex-stratified randomization (**Table S2-Now-New-Fig**). Across acute response time points (3h and 24h after exercise in muscle), the response was generally greater at 24h compared to 3h for both W0 and W12 (**Figure 3A**). At 24h post-exercise, the number of DE miRs appeared higher in TRAD at W0 (20 vs. 13) but higher in HITT at W12 (32 vs. 11), indicating distinct response patterns between the exercise doses. DE miRs were largely unique to TRAD or HITT with limited overlap (**Figure 3A**, “shared”). However, five DE miRs in the 24h acute response were shared across doses and training state (miR-7-5p, miR-142-3p, miR-223-3p, miR-3529-3p and miR-582-5p). These shared miRs appear to represent a general exercise response in muscle that persisted even after 12wk of training (**Table S2-Now-New-Fig**). Notably, previous studies have identified miR-223-3p as responsive in both serum EVs and muscle and miR-582-5p as responsive to a single exercise bout in the untrained state. Among the shared miRs, miR-3529-3p is unique in having no reported link to exercise (11).

**Figure 3.**
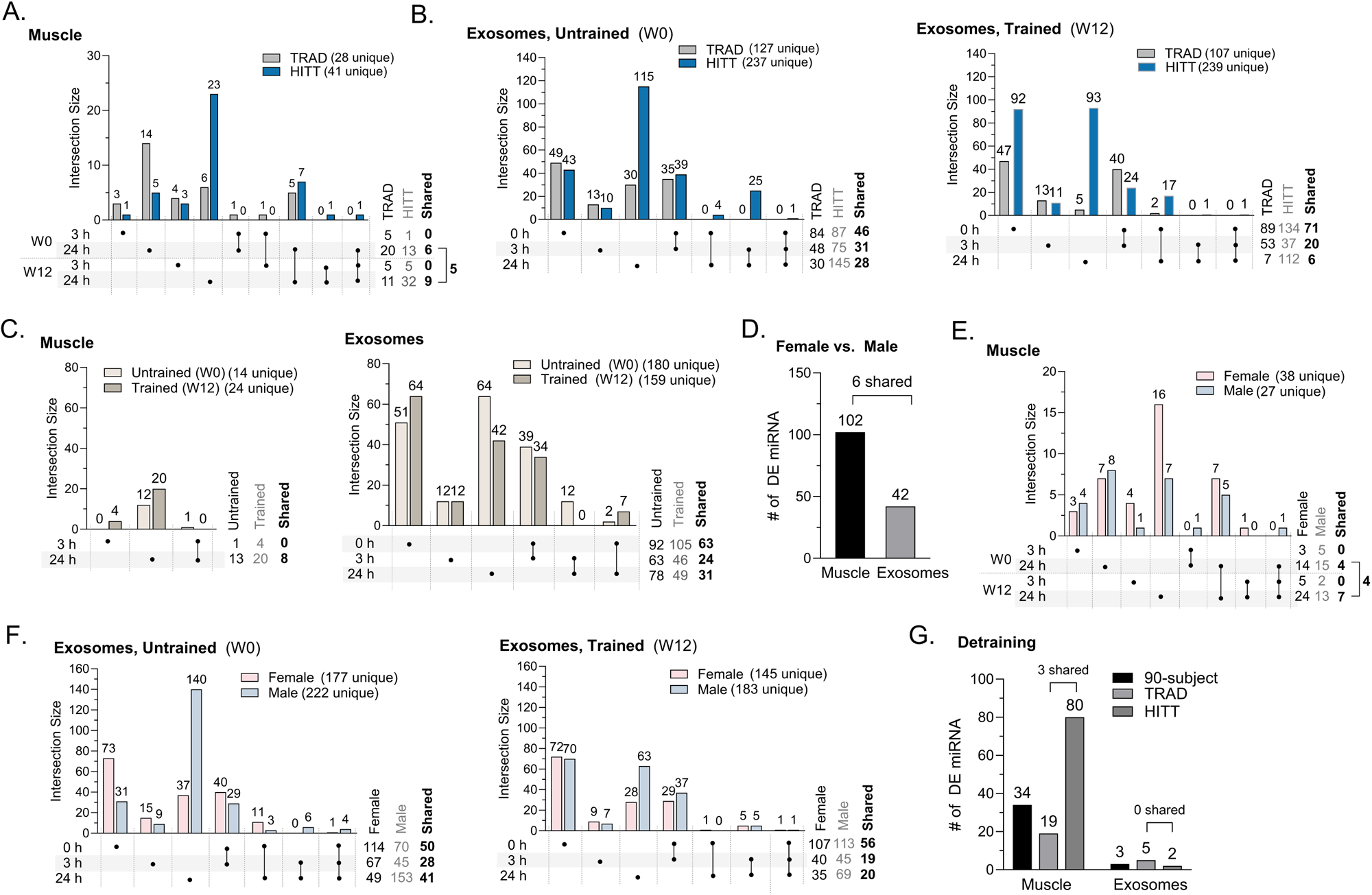
MiR signatures of exercise prescription, training status and sex are evident in serum exosomes and muscle. Upset plots (A,B,C,E,F) show the number of miR differentially expressed (DE) versus the corresponding baseline (w0 or w12 baseline sample). DE miR are identified as those with p≤0.05, │FC│≥1.5, and a minimum average read count of 10 reads in each category. Comparisons between 3h and 24h (muscle) or 0h, 3h and 24h (exosomes), and/or between untrained (W0) and trained (W12) states are shown. Numbers above each bar indicate the number of DE miR, and numbers to the right indicate the total DE counts by category and shared between categories. (A-C) Upset plots comparing miR DE in each exercise prescription separately (n=43 TRAD or n=47 HITT) for muscle (A) and exosomes (B, W0 on left, W12 on right) or when analyzed together (C) to ascertain miRs DE in the general response to exercise (n=90) in muscle (left) and exosomes (right). (D) DE miR at baseline between Females (n=48) and Males (n=42). (E,F) UpSet plots comparing miR DE in muscle in response exercise in Females (n=48) and Males (n=42) in muscle (E) and exosomes (F) of untrained (left) and trained (right) participants. (G) DE miR associated with detraining (W16 vs W12 pre) in muscle (n=89) and exosomes (n=89) for the overall response and for TRAD and HITT separately.

### Serum exosomes

In serum exosomes, analysis of pre-exercise baseline in TRAD vs. HITT yielded only 9 DE miR (**Table S2-Now-New-Fig**). A marked early response in serum exosome miR levels was observed immediately post-exercise (0h) in both untrained and trained states (**Figure B**). Only one miR, miR-197-3p, was DE at all time points (0h, 3h, 24h) in both W0 and W12, and it was specific to the HITT group. miR-197-3p is a known exercise-responsive miR, known to be upregulated in plasma after a single bout of endurance or resistance exercise (32, 33). In TRAD, 127 and 107 unique miRs were DE at W0 and 12 (across 0, 3 or 24h), respectively with 59 DE miRs common to both time points. In HITT, 237 and 239 unique miRs were DE at W0 and W12, respectively (across 0, 3 or 24h) with 172 DE miRs common to both W0 and W12 (**Figure 3B**, **Table S2-Now-New-Fig**). Among the DE miRs, 42 miRs were shared between TRAD and HITT. Of note, two of these shared miRs, miR-223-3p and miR-582-5p, were DE in both exosomes and muscle (**Figure 3A, 3B**, **Table S2-Now-New-Fig**). Both miRs are known to be altered in response to exercise. Specifically, miR-223-3p was DE in serum-derived EVs 24h after a single exercise bout in the untrained state (11); Previous studies have also reported its up-regulation in serum after marathon running and DE in serum after ∼6 months of training (34, 35). miR-582-5p is downregulated in EVs after fatiguing freestyle swimming (36–38).

### Training state (untrained, trained) influenced overall acute exercise response miR signatures irrespective of exercise prescription

Since *in vivo* physiological performance and body composition adaptations were largely unaffected by exercise prescription (**Table 1**), we further examined the impact of training state on miR signatures in serum exosomes and muscle in the full cohort (n = 90). In muscle, a robust response was noted at 24h at both W0 (13 of 14 DE miRs) and W12 (20 of 24 DE miRs) (**Figure 3C, left**). Among the 38 total unique miRs DE in muscle, only miR-223-3p was shared between W0 (DE at 3h and 24h) and W12 (DE at 24h).

Consistent with the prescription-specific analysis, the magnitude of response to exercise was greater in serum exosomes than in muscle. Extensive changes in exosomal miR levels were observed at 0h in both untrained (180 DE miRs) and trained (159 DE miRs) states (**Figure 3C, right**), collapsed across 0, 3 and 24h. Of these, 108 DE miRs were shared between untrained and trained states (**Figure 3C, right**), resulting in 231 unique exosomal miRs DE post-exercise. Forty-two of these 108 miRs were identical to the 42 shared between TRAD and HITT when analyzed separately (Figures 2B-C and Table S2-Now-New-Fig). Of the 28 muscle miRs and 231 exosomal miRs altered with exercise, 12 miRs were shared across muscle and exosomes (**Figure 3C**). Of these, six are newly discovered exercise-responsive miRs (miR-181c-5p, miR-514a-3p, miR-3680-5p, miR-4684-3p, miR-4714-5p and miR-6082). The remaining six are well-characterized exercise responsive miRs showing DE in multiple tissue types (miR-142-5p, miR-146b-5p, miR-206, miR-223-3p, miR-424-5p and miR-582-5p) (**Table S2-Now-New-Fig**) (36, 38–44).

### Exercise-responsive miRs target mRNAs in known exercise-related pathways

High confidence mRNA targets of DE miRs, identified via miRTarbase v9, were subjected to KEGG pathway enrichment analysis using Enrichr to identify predicted signaling pathways altered in response to exercise. For the muscle acute response, we combined all muscle miRs DE in response to exercise (i.e. 3h and/or 24h post-exercise at W0 and/or W12; **Figure 3C, left**). As expected, high-confidence mRNA targets showed enrichment in typical exercise-related pathways including mTOR signaling (odds ratio = 8.56, combined score = 280.15, adjusted p = 2.57E-14) and AMPK signaling (odds ratio = 7.32, combined score = 154.24, adjusted p = 2.04E-09). Similarly, high-confidence mRNA targets of DE exosomal miR (**Figure 3C, right**) showed enrichment in mTOR (odds ratio = 10.25, combined score = 638.61, adjusted p = 5.53E-27) and AMPK (odds ratio = 9.18, combined score = 398.22, adjusted p = 5.80E-19) signaling.

We further analyzed muscle responses separately in untrained (**Figure S3A, Table S4-Now-New-Fig**) and trained (**Figure S3B, Table S4-Now-New-Fig**) states. High confidence mRNA targets in the untrained state mapped to multiple metabolic and pro-inflammatory pathways (**Figure S3A**), as well as pathways involved in cell cycle regulation (e.g., p53, PI3K-AKT, Hippo), capillary angiogenesis (HIF-1), and autophagy. Interestingly, the trained state acute response mapped to largely the same pathways (**Figure S3B, Table S4-Now-New-Fig**). Among miRs with no prior known association to exercise responses, we identified miR-524-5p in the untrained state and 3 novel miRs in the trained state (miR-181c-5p, miR-183-5p and miR-421).

### Sex differences are profound in miR signatures at baseline and following exercise

Sex differences were tested in serum exosome and muscle samples from W0 pre-exercise (fasted, morning, resting) in females and males. A total of 102 muscle and 42 exosomal miRs were DE between sexes (**Figure 3D**). This finding is consistent with previously reported sex differences in miR profiles in serum, as well as gonadal and nongonadal tissues (45–48). Among these, six miRs (miR-15b-5p, miR-199a-3p, miR-199b-3p, miR-361-5p, miR-381-3p and miR-12116) differed by sex in both muscle and exosomes. Five have been previously linked to exercise and shown to differ between sexes (36, 38–42, 44, 49). However, miR-12116 has not been previously associated with exercise or sex differences.

Next, we explored sex differences in the acute miR responses to exercise. In muscle, the magnitude of the exercise response was similar between sexes (**Figure 3E, Table S2-Now-New-Fig**) and was most pronounced at 24h in both untrained (W0) and trained (W12) states. While a few DE muscle miRs at 24h were shared by both sexes, the majority exhibited sex-dependent responses (**Figure 3E**). Four acute response DE miRs shared between sexes in both untrained and trained states (miR-142-3p, miR-223-3p, miR-3529-3p and miR-7-5p) (**Figure 3E, Table S2-Now-New-Fig**) were also identified in the analysis of the combined sex cohort, suggesting a role for these miRs in the general muscle exercise response, independent of sex or training state.

In serum exosomes (**Figure 3F**), untrained females showed a robust acute exercise response immediately post-exercise (0h) with 114 DE miRs, while in untrained males the dominant response occurred at 24h post-exercise, with 153 DE miRs (**Figure 3F, Table S2-Now-New-Fig**). In the trained state, there were no notable sex differences in the timing of acute exercise responses (**Figure 3F**). Fewer than half of the acute DE miR responses were shared across the sexes (**Figure 3F**, “shared”). Only miR-197-3p was DE at all time points (0h, 3h, 24h) at W0 and W12 and exclusively in males. Notably, only miR-223-3p exhibited differential regulation in both serum exosomes and muscle (**Figure 3E and 3F**, **Table S2-Now-New-Fig**).

## DETRAINING

### Short-term detraining reversed some but not all ex vivo muscle tissue adaptations

Detraining induced myofiber atrophy was prevalent in HITT group as noted for type II and mean fCSA while no significant atrophy was observed in the TRAD group, **Table 2**). However, training-induced improvements in capillarization were retained in both TRAD and HITT, indicating relative short-term stability (**Table 2**). Conversely, training-induced increases in Pax7^+^ SCs and indices of mitochondrial biogenesis were completely reversed after 4wk of detraining period (**Table 2)**.

### A detraining miR signature is evident in muscle but not in serum exosomes

A significant detraining signature was evident in muscle with 34 DE miRs (**Figure 3G**), 28 of which have known links to exercise (**Table S2-Now-New-Fig**). Within-group follow up analysis revealed a more pronounced detraining signature in HITT (n=80) vs. TRAD (n=19), with minimal overlap (n=3) between the two prescriptions (**Figure 3G**). A robust network of high confidence mRNA targets was observed for miRs in the detraining muscle signature (**Figure S3C**). Among these, three miRs (miR-148a-5p, miR-181C-5p and 450a-5p) had no prior linkage to exercise or detraining. Pathway Enrichment analysis (**Figure S3C**) for detraining muscle showed many of the same pathways found in the untrained (**Figure S3A**) and trained (**Figure S**3**B**) exercise responses, but with far greater transcript membership. Pathways unique to detraining included TNF, Ras, and Toll-like receptor signaling, indicating potential alterations in inflammation, cell survival, remodeling, and immune signaling in the early phase of muscle detraining. In contrast to muscle, detraining appeared to exert little impact on exosomal miR (**Fig. 3G, Table S2-Now-New-Fig and S4-NOW-NEW-FIG**).

## INTER-INDIVIDUAL RESPONSE HETEROGENEITY

### In vivo adaptations to training vary widely among individuals and are independent of exercise prescription

There has been a growing focus on understanding the significant variation in the individual response to exercise, with the goal of developing personalized exercise prescriptions to maximize health benefits and optimize individualized rehabilitation (50, 51). The combined training programs in this trial provided a unique opportunity to investigate inter-individual response heterogeneity (IRH) across clinically important indices of both endurance training adaptability [cardiorespiratory fitness (CRF), i.e., VO_2_peak] and resistance training adaptability [functional muscle quality (fMQ), i.e., knee extensor strength normalized to DXA-derived thigh lean mass]. We applied a well-established distribution-based method (27) to define minimum clinically important difference (MCID) scores pre- to post-training in CRF and fMQ. Eighty-two participants were classified based on these MCID criteria shown in **Figures 4A, 4B**. Importantly, the proportions of participants attaining or exceeding these MCIDs were not biased by training prescription (HITT, TRAD) (**Figures 4A, 4B**), allowing us to pool all participants as one cohort to better interrogate IRH. Overall, 67% of participants attained the MCID for fMQ while 61% attained the CRF MCID. The distribution of participants achieving both MCIDs (39%), fMQ only (28%), CRF only (22%), or neither (11%) is summarized in the Punnett square (**Figure 4C**). From the cohort of 82 participants with MCID classification, we downselected n=80 for DGE and n=75 for higher-order singular value decomposition (HOSVD) due to missing features needed for each analysis.

**Figure 4.**
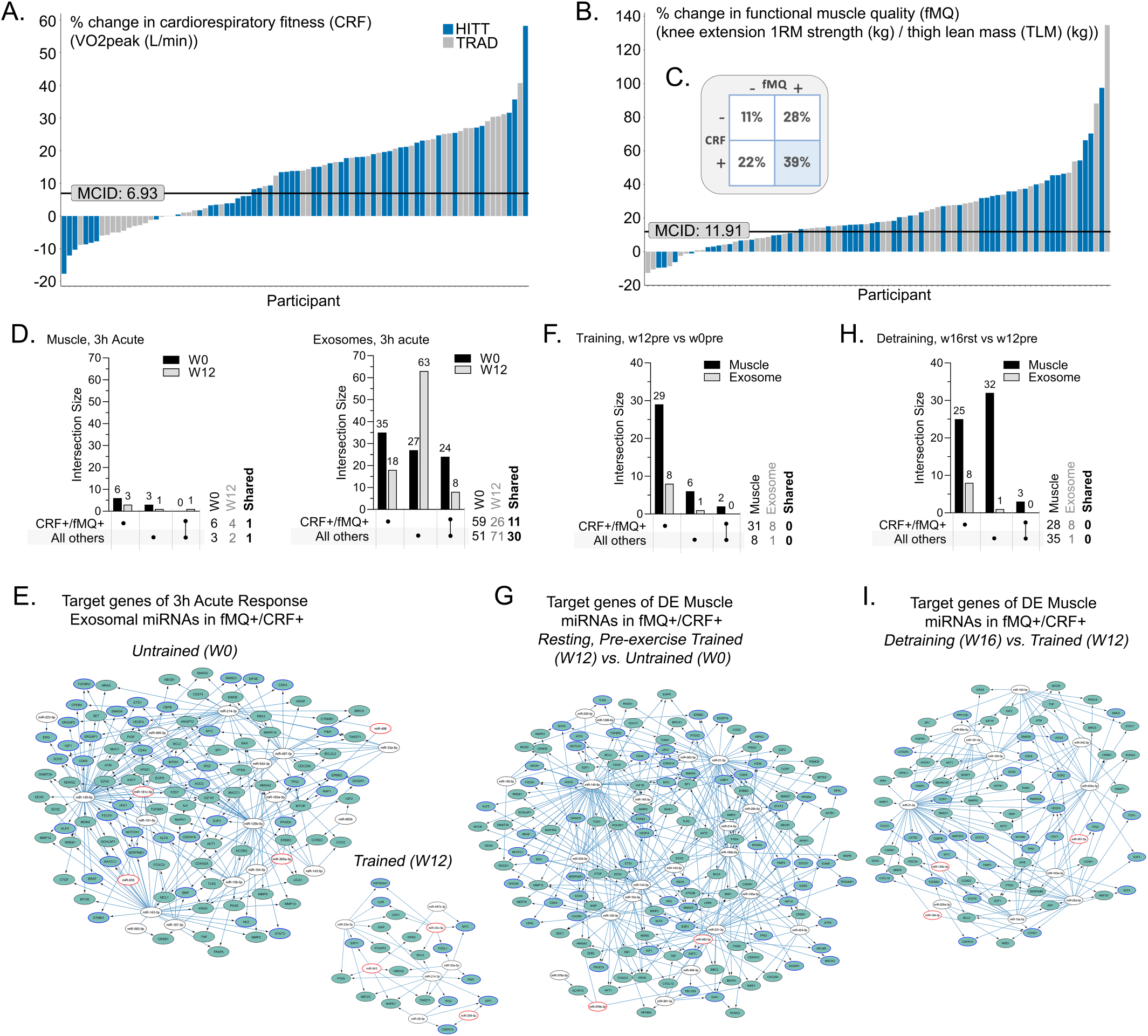
Wide inter-individual response heterogeneity in clinically important outcomes and linked miR signatures. (A-B) Waterfall plots showing individual’s % change in cardiorespiratory fitness (CRF) (A) and % change in functional muscle quality (fMQ) (B). Individual responses are color coded for HITT and TRAD participants. Horizontal line is the minimum clinically important difference (MCID). (C) Punnett square classification of participants who attained (+) or did not attain (-) MCID for fMQ and CRF. (D-I) Analysis of differentially expressed (DE) miR in the acute response (D-E; 3hr versus baseline pre sample), training (F-G; W12pre vs w0pre) and detraining (H-I; w16rst vs w12pre) in fMQ+/CRF+ dual responders (n=30) and all others (n=50) in muscle and exosomes, as indicated. Upset plots (D,F,H) detail DE miR identified as those with p≤0.05, │FC│≥1.5, and a minimum average read count of 10 reads in each category. Numbers above each bar indicate the number of DE miR, and numbers to the right indicate the total DE counts by category and shared between categories. The corresponding target gene networks (E,G,I) show high-confidence mRNA targets regulated by at least 2 DE miR in the dataset. miR with red outlines have no known link to exercise. mRNA nodes with blue outlines are exercise-responsive mRNAs in MetaMEx. High confidence Cytoscape gene networks are available as NDEX libraries (with links provided in Table S3-Now-New-Fig).

### miRs differentially regulated in fMQ+/CRF+ target mRNAs in exercise related signaling pathways

The miR signatures of fMQ+/CRF+ dual responders (n=30) were compared to the remaining cohort (n=50). DGE analysis revealed a more pronounced acute (3h) exercise response in exosomes compared to muscle (**Figure 4D, left and Table S2-Now-New-Fig**), with the fMQ+/CRF+ participants showing a relatively blunted trained response (W12). Among fMQ+/CRF+, 11 DE miRs were shared across the W0 and W12 exosome responses compared to 30 DE miRs among the other participants across these timepoints (**Figure 4D, right**). Cytoscape analysis revealed exosomal miR links to exercise which have not been previously reported. These include miR-181c-5p, miR-365a-3p, miR-496 and miR-935 in untrained acute response, and miR-34c-5p, miR-299-5p and miR-543 in the trained acute response (**Figure 4E**). No DE miRs were shared between muscle and exosomes. In contrast to the acute responses, the effect of training (*i.e.*, W12 baseline trained state vs. W0 baseline) was more evident in muscle, with more DE miRs in fMQ+/CRF+ vs. the others (31 and 8, respectively) (**Figure 4F**). The Cytoscape network of DE miRs from muscle in the fMQ+/CRF+ group included two not previously linked to exercise, miR-376b-3p and miR-483-3p (**Figure 4G**). Similarly, the effect of detraining was more dominant in muscle, with no overlap in signatures between muscle and exosomes (**Figure 4H**). The fMQ+/CRF+ detraining Cytoscape network from muscle included three miRs not previously linked to exercise, miR-135b-3p, miR-194-3p and miR-361-5p (**Figure 4I**). Pathway analysis of the high confidence mRNA targets of the DE miRs from signaling networks in **Figure 4** indicated enrichment in MAPK, PI3K-AKT, HIF-1, mTOR, and cell cycle regulatory pathways, among others (**Figure S4, Table S4-Now-New-Fig**). Interestingly, longevity regulating pathway signaling was enriched in the acute and trained response for fMQ+/CRF+ participants but not in detraining.

### HOSVD analysis reveals in vivo and ex vivo phenomic features associated with fMQ+/CRF+ responder status

We leveraged HOSVD to define multidimensional biocircuitry by integrating all phenomic, ex vivo, and molecular features (**Figure 5**) linked to the dual responder fMQ+/CRF+ classification (**Figure 5A**, Punnett sub-square shaded blue) versus all others (**Figure 5A** Punnett sub-squares shaded white). Responder status was not biased by sex for CRF+/- or fMQ+/- nor did it appear as an important feature for fMQ+/CRF+. The HOSVD model incorporated over 37,000 features across the categories in **Figure 5B**. Top phenomic features are summarized in **Figure 5C** (directionality of each arrow indicates fMQ+/CRF+ vs. all others). Interestingly, the majority linked to MCID attainment were baseline pre-training features. At baseline, fMQ+/CRF+ group reported lower physical activity, with 58% identifying as sedentary compared to 29% in other groups. Additionally, 81% of the fMQ+/CRF+ participants reported low workplace physical demands, compared to vs. 59% in the remaining cohort. Habitual caffeine (coffee, tea, soda) consumption was lower as 58% of fMQ+/CRF+ reported none vs. 35% in non-fMQ+/CRF+.

**Figure 5.**
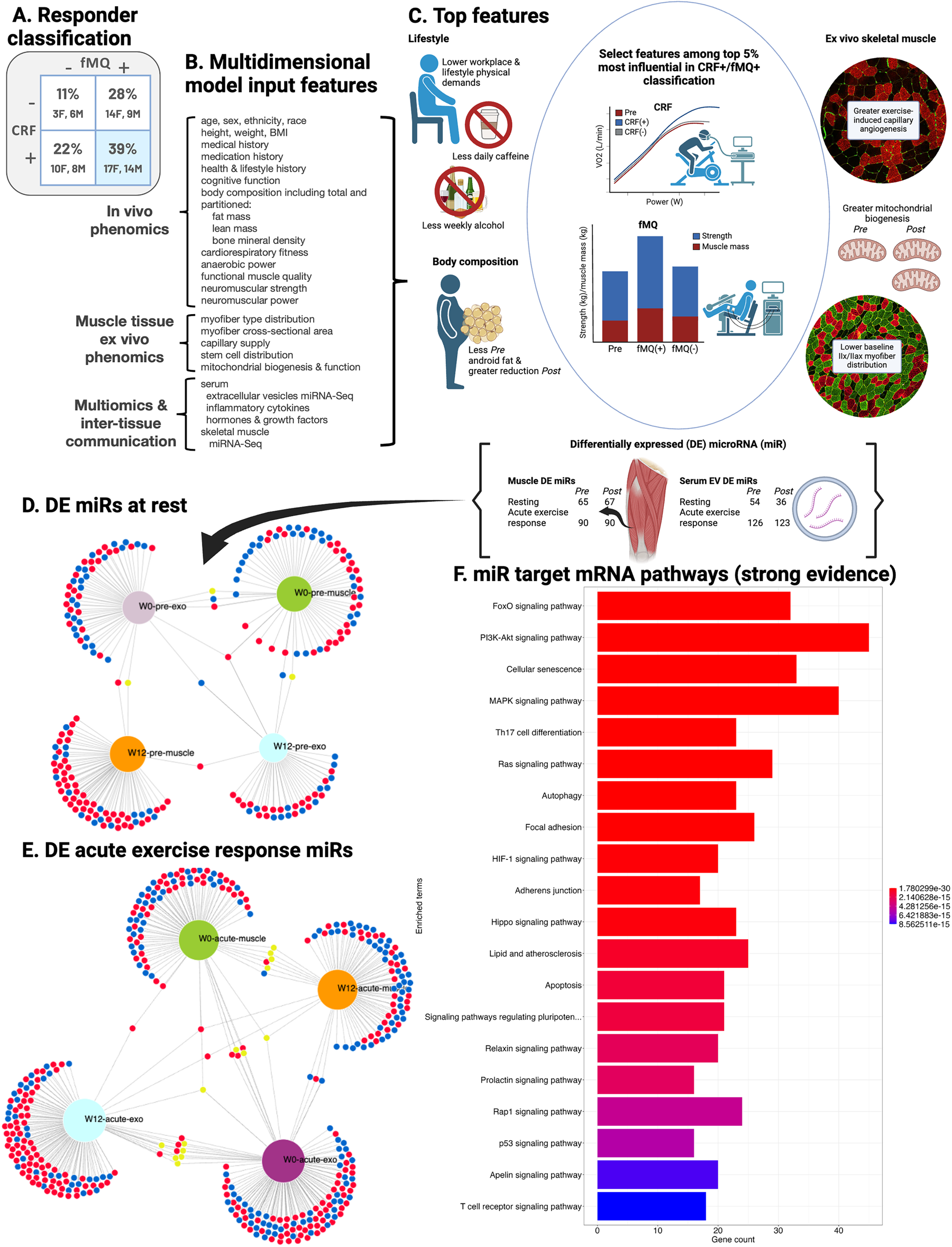
Multidimensional biocircuitry linked to inter-individual response heterogeneity via higher order singular value decomposition (HOSVD). (A) Punnett square displaying the observed distribution of minimum clinically important difference (MCID) scores for functional muscle quality (fMQ) and cardiorespiratory fitness (CRF); achieved MCID (+); did not achieve MCID (-). Distribution by sex is also shown. (B) Categories of HOSVD input features to model biocircuitry of fMQ+/CRF+ dual responders vs. all others. The input features across these categories totaled 881 phenomic and 37,026 miR (including all timepoint contrasts in exosomes and muscle). (C) Summary of key input features among the down-selected most influential top 5% of features from the HOSVD models of MCID classification fMQ+/CRF+ vs. all others. (D) DiVenn plot of muscle and exosomal differentially expressed miRs (from top 5% of HOSVD features) in fMQ+/CRF+ vs. all others in fasted, morning resting samples (prior to the acute exercise bout) in the untrained state (W0) and trained state (W12). Red = upregulated, blue = downregulated, and yellow = miR shared across nodes that are upregulated in one node and down in another. (E) DiVenn plot of muscle and exosomal differentially expressed miRs (from top 5% of HOSVD features) in fMQ+/CRF+ vs. all others collapsed across the acute exercise response time points for serum exosomes (0h, 3h, 24h) and for muscle (3h, 24h). Red = upregulated, blue = downregulated, and yellow = miR shared across nodes that are upregulated in one node and down in another. Acute response miRs were determined as change from pre-exercise within the given acute response bout (i.e., untrained W0 or trained W12). (F) KEGG 2021 top 20 non-disease enriched pathways (p≤0.05) resulting from downselection of the differentially expressed pre-exercise and acute response miRs at W0 and W12 in serum exosomes and muscle with high confidence (strong evidence) mRNA targets.

Habitual alcohol intake was also lower in fMQ+/CRF+ with 46% reporting no regular consumption whereas 33% of non-fMQ+/CRF+. Android adiposity was an important component as trunk fat mass was 1.6kg lower in fMQ+/CRF+ vs. non-fMQ+/CRF+ with various components of android adiposity also being lower in fMQ+/CRF+ such as trunk subcutaneous adipose mass (0.274kg lower) and arm fat mass (.290kg lower). Android adiposity was also the dominant training-induced *in vivo* phenotype difference between fMQ+/CRF+ and all others as fMQ+/CRF+ had nearly double the decrease in trunk (9% vs. 5%) and android percent fat (11% vs. 6%) compared to non-fMQ+/CRF+. Android fat was a key phenotypic feature at both baseline and change across exercise training. No overt inflammation was detected, however circulating IL-6 levels were slightly lower in fMQ+/CRF+ (∼0.122 pg/mL). Via muscle tissue analyses ex vivo, fMQ+/CRF+ had lower baseline muscle capillary density (246 vs. 268 cap/mm^2^) and fewer capillary contacts per type I myofiber (3.84 vs. 4.08). In muscle, capillarization increased to a greater extent in fMQ+/CRF+ which may reflect the lower starting point.

### HOSVD analysis reveals a differential resting and acute response miR expression profiles associated with fMQ+/CRF+

Downselection of the top 5% of features in the top 10 LVs identified 524 miR features highly associated with fMQ+/CRF+ (**Figures 5D and 5E**), with 236 DE miRs noted in muscle alone, 258 in serum exosomes alone, and 30 shared between tissues. **Figure 5D** highlights the 222 DE miRs (132 in muscle, 90 in exosomes) detected under resting conditions (fasted, morning pre-exercise at W0 or W12). In resting muscle, 65 DE miRs were detected at W0 with 67 at W12 (overall, 50 higher and 82 lower in fMQ+/CRF+). Among the 90 exosomal DE miRs, 54 were detected at W0 and 36 at W12 (overall, 43 were higher and 47 lower in fMQ+/CRF+). Minimal overlap in DE miRs was observed between tissues or between W0 and W12 resting timepoints (**Figure 5D**). Four were shared between muscle and exosomes at W0 and only one at W12. **Figure 5E** summarizes the miR profiles differentially responsive to acute exercise in fMQ+/CRF+ (vs. all others) at both W0 and W12. Across acute response time points (0h, 3h, 24h) at W0 and W12 HOSVD and subsequent downselection of features retained 447 DE miRs with 94 at W0 and 93 at W12 in muscle (39/94 and 55/93 higher in fMQ+/CRF+) and 131 at W0 and 129 at W12 in exosomes (55/131 and 39/129 higher in fMQ+/CRF+). KEGG pathways analysis was performed on high confidence target mRNAs (**Figure 5F and Table S4).** The resting and acute response profile of fMQ+/CRF+ (vs. all others) suggests differential regulation of several pathways central to stem cell cycle regulation (*e.g.*, Signaling pathways regulating pluripotency of stem cells, p53, Ras), autophagy, and apoptosis (**Figure 5F, Table S4**). Additionally, pathways that regulate processes favorable to positive physiologic adaptations were also altered, including blood flow, angiogenesis, endothelial function (e.g., relaxin, HIF-1, MAPK), and metabolism (PI3K-AKT, MAPK, FoxO, **Figure 5F, Table S4**).

### Integration of DGE and HOSVD pipelines reveals nine miRs that may be central to exercise adaptability

To further our understanding of the miR-mediated modulation of individual adaptability to exercise training (*i.e.*, IRH) we compared the select miRs retained after strict down-selection processes in the HOSVD modeling framework to the DE miR signatures obtained through straightforward DE analysis of the miR data alone in the context of MCID attainment. We constricted this cross-comparison to: (i) 3h acute responses in the untrained state (W0); (ii) 3h acute responses in the trained state (W12); and (iii) resting expression profiles at W12 vs. W0 (i.e., training responses). A total of 168 unique miRs were identified by DGE analysis and 59 by HOSVD, with 9 common to both pipelines (**Figure 6A, Table S2-Now-New-Fig**). Five of the nine miRs have no known prior linkage to exercise (**Figure 6B**). The high confidence mRNA targets are largely unique to each of the nine miRs with only two target genes shared (BMI1, ERBB2) between two miRs (**Figure 6B**). Pathway analysis of these high confidence mRNA targets (**Figure 6C, Table S4-Now-New-Fig**) showed enrichment in FoxO, JAK-STAT, HIF-1, TGF-β, and PI3-AKT signaling, along with stem cell regulation (among others).

**Figure 6.**
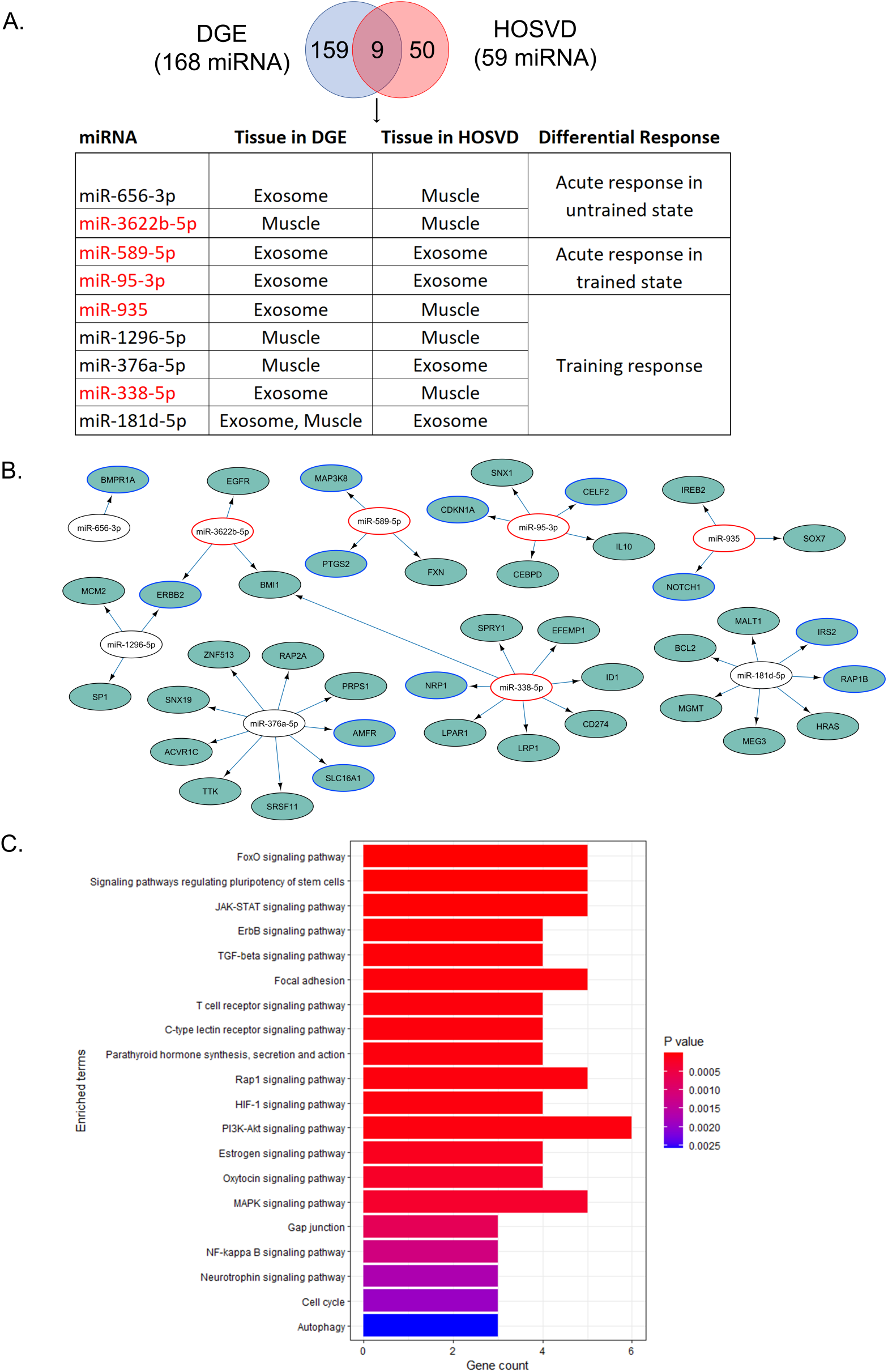
Cross-validation of miRs linked to inter-individual response heterogeneity. The cross-validation was restricted to: (i) 3h acute responses in the untrained state (W0); (ii) 3h acute responses in the trained state (W12); and (iii) resting expression profiles at W12 vs. W0 (i.e., training responses). (A) Venn diagram comparing the miR resulting from these analyses (n=168 unique miRs identified using differential gene expression (DGE) analysisand n=59 miRs from the top 5% of HOSVD features with nine common to both pipelines). The 9 miR common to both pipelines are featured in the table. Five of the nine miRs (indicated by red text) have no known prior linkage to exercise. (B) Target gene networks of DE miR in fMQ+/CRF+ responders generated from the nine miRs common to both DE and HSOVD pipelines after filtering to only high-confidence mRNA targets. miRs with red outlines are miRs with no known link to exercise. mRNA nodes with blue outlines are exercise-responsive mRNA in MetaMEx. (C) KEGG 2021 top 20 non-disease enriched pathways (p≤0.05) of the high-confidence mRNA targets from panel B.

## DISCUSSION

Long-term exercise training is the only lifestyle choice to date that has consistently demonstrated an ability to improve function and health across multiple tissues and organ systems. While single mode exercise prescriptions (*e.g.,* MICT, HIIT or progressive RT) improve specific indices of physiological function and overall health with some cross-over, adaptations to combined training have received much less attention.

### HITT vs. TRAD phenomic adaptations

We show here that a 3 d/wk HITT prescription combined with 2 d/wk high-intensity, short rest interval RT is as efficacious as a TRAD regimen despite being less than half the duration per session. HITT equaled TRAD for gains in aerobic power as measured by VO_2_peak and both TRAD and HITT induced central and peripheral adaptations resulting in training-induced gains in VO_2_peak. Interestingly, our findings suggest TRAD was advantageous for enhancing the mechanisms driving both O_2_ pulse and capillary angiogenesis, while HITT more effectively induced biogenesis of IMF mitochondria, the pool of myofiber mitochondria typically resistant to exercise adaptation (**52**).

Even though the RT prescriptions in HITT and TRAD differed substantially in loading intensity (9 RM vs. 13 RM) and work/rest intervals, gains in strength and muscle power were generally comparable except for upper body strength gains which favored HITT. The nearly two times greater gain in anaerobic power among HITT compared to TRAD was not surprising given the principle of training specificity: the HITT prescription involved two Wingate-like 30s maximal power cycle sprints during each HITT session 3 days per week (*i.e.*, 2 of 10 HITT intervals each session). Although gains in total lean mass and limb lean mass values did not differ by prescription, hypertrophy at the myofiber level was superior in TRAD and this advantage was associated with a more robust increase in the prevalence of Pax7^+^ SCs.

While a few phenotypic adaptations favored TRAD or HITT, the findings overall indicate the two prescriptions were similarly efficacious at inducing core health benefits with established clinical importance (*e.g.*, CRF, leg strength, muscle mass). It would be valuable to determine whether the peripheral adaptation advantages (e.g., myofiber hypertrophy and capillarization) seen here in TRAD result in continued, progressive gains or a divergence of performance adaptations over longer term training.

### Detraining signature in muscle

After four weeks of detraining, indices of muscle mitochondrial capacity returned to baseline levels, in agreement with previous literature (**53, 54**). Training-induced gains in muscle capillarization remained after 4 weeks of detraining, consistent with prior work (**53**), although others have reported partial reversal during a similar detraining period.(**55**) The detraining-induced reduction in the histological prevalence of Pax7+ muscle SCs is also consistent with previous studies in young (**56**) and older (**57**) adults. Though detraining leads to reversal it should not be assumed that the effects of exercise are lost. Previous work suggests that a “memory” signature exists in muscle stem cells that facilitates adaptation when exercise is resumed (**58**). In older adults it has been shown that if exercise training is resumed following detraining, SC abundance may rebound even above the increase from the previous training period (**57**).

Interestingly, detraining myofiber atrophy occurred in an exercise prescription-specific manner. TRAD participants retained the training-induced hypertrophy noted for type II myofibers and mean fCSA, whereas HITT lost all gains in myofiber size. Myofiber hypertrophy, which occurs with endurance training (59–61), especially with cycling (61, 62) in untrained adults, appears to be retained during short-term detraining (63, 64). The preserved fCSA unique to TRAD may therefore have been linked to mechanisms unique to MICT and the cycling modality used. Detraining DE miRs in HITT mapped to 126 of the 354 mRNAs in the KEGG PI3K-AKT signaling pathway vs. only 12/354 for TRAD, suggesting downstream mTOR-mediated protein synthesis signaling (65) was differentially impacted in HITT versus TRAD during detraining. Considering the typical inhibitory nature of miRs, this suggests greater suppression of protein synthesis in HITT, leading to the detraining atrophy seen in HITT but not TRAD.

### Differentially expressed miRs across serum exosomes and muscle

Over 1,000 miRs have been linked to various exercise modalities, durations, intervals, intensities and sample types. (Table S2-Now-New-Fig lists PMIDs linking miRs in this study to exercise with relevant sample types indicated. *The full manually curated list of exercise-responsive miR is available upon request*). Since the majority of exosomal miRs originate from vascular endothelial cells, platelets, lymphocytes and monocytes (**66**), it follows that the miR signatures of the exercise response in serum exosomes and muscle would be largely independent. Consistent with this prediction and prior reports (**67, 68**), our observation that the serum exosomal miRs altered in response to exercise were mostly distinct from those altered by exercise in muscle implies that miR release from muscle is not a dominant contributor to subsequent changes in circulating miR levels.

We observed significant baseline (*i.e.*, pre-exercise) differences in the levels of 102 muscle-derived miR and 42 serum exosomal miRs between females and males (**Figure 3F**). This finding is in line with published reports indicating that the acute and chronic integrative molecular response to exercise differs between males and females (69, 70). Six of the miRs DE between males and females at baseline were found to be shared across muscle and exosomes (miR-15b-5p, 199a-3p, 199b-3p, 361-5p, 381-5p and miR-12116). All six were found to be DE in response to exercise in at least one of the comparisons (**Figure 3**). Since the study population overall and within HITT and TRAD was comprised of roughly equal numbers of males and females, all 6 are presumably exercise responsive miRs with “sexually dimorphic” expression.

We observed a robust DE response among serum exosomal miRs immediately post-exercise in both untrained and trained states while the DE response for muscle-derived miRs peaked 24h post-exercise. These response kinetics mirror previous reports indicating changes in serum exosomal miR expression immediately after cycling exercise with a decline to baseline levels within 90 minutes of rest (71), and an upregulation of muscle miRs peaking around 8h post-exercise (72). The effects of single mode exercise interventions on muscle and exosomal miR levels have largely focused on the acute (i.e. <24h) response (73–75), while the effects of extended training and/or detraining have been largely ignored. Of the 12 miRs associated with a 6-month detraining period in mice (58), none linked to detraining in this trial. Our results suggest a much more potent miR response to detraining in muscle than in serum exosomes.

### Multidimensional modeling and integration

The key purpose of employing HOSVD was to map biocircuits of IRH integrating both phenomic and miR signatures that converge on the fMQ+/CRF+ classification. The dominance of miR among the down-selected features (524 miR vs. 87 phenomic) is reflective of the overall numbers of input features (37,026 miR vs. 881 phenomic); not an indication of preferential importance for miRs. The existence of integrated circuits associated with MCID attainment (i.e., fMQ+/CRF+ vs. all others) is supported by the identification of LVs related to inflammation, angiogenesis, and adiposity. IL-6, which promotes cell senescence and tissue remodeling in healthy individuals (**76**) but is associated with disease and maladaptation when chronically elevated (**77**), was lower in fMQ+/CRF+ individuals at baseline. There were also overlapping signatures between capillary supply and miR targeted pathways related to angiogenesis. HOSVD predicted changes in the PI3K/AKT pathway, a potent stimulator of angiogenesis and endothelial cell proliferation (**78, 79**), and HIF-1 signaling, a key regulator of angiogenesis and endothelial cell function (**80**) as well as stem cell function. Trunk fat, specifically android fat, was lower in fMQ+/CRF+ and there was greater android fat loss in fMQ+/CRF+ across training. FOXO signaling which is a key promoter of fat oxidation and mitochondrial lipid metabolism was differentially regulated in fMQ+/CRF+ (**81**). It is important to note that only 131/524 had strong evidence for miR/gene target interaction and were used for pathway analysis. Thus, much remains unknown about the pathways linked to fMQ+/CRF+ responder status.

### Nine miRs linked to fMQ+/CRF+ are robust to methodological and sampling differences of DGE and HOSVD

In addition to the mathematical differences in the DGE and HOSVD approaches, the pipelines differed in number of participants (80 in DGE analysis of MCID categories vs. 75 in HOSVD), miR data format (i.e., individual timepoints in DGE versus change scores between timepoints in HOSVD) and inclusion of metadata (i.e. sample type only in DGE analysis by MCID, vs. extensive metadata use in HOSVD). The approach taken here serves as an illustration of the power of combining multi-dimensional models with standard DE pipelines as a means of prioritizing and identifying influential features. The fact that the same nine miRs appear in the output of both pipelines is evidence of the robustness of the association of these miRs with individual attainment of clinically important exercise-induced health benefits (e.g., fMQ and CRF).

Among the nine miRs, miR-656-3p and miR-3622-5p were linked to the acute exercise response in untrained individuals. miR-656-3p is downregulated in the plasma of female synchronized swimmers (82), promotes viability and migration of placental mesenchymal stem cells, and targets HIF-1alpha (83), suggesting a potential role in exercise-induced tissue hypoxia and angiogenesis. Although miR-3622b-5p has no reported link to exercise, it targets ERBB2 which plays a role in skeletal muscle repair and fibrosis in rats (84).

miR-589-5p and miR-95-3p had no previous link to exercise that we found acutely responsive to exercise in the trained state. miR-589-5p, which was upregulated in exosomes of the nonresponders in the trained state, is reported to reduce oxidative stress in human retinal microvascular endothelial cells via targeting EGR1 (85), suggesting a possible response to oxidative stress in nonresponders. miR-95-3p has been shown to regulate cyclin-dependent kinase inhibitor 1A (CDKN1A), suggesting a role for miR-95-3p in the increased levels of CDKN1A reported to occur after exercise (86, 87) and the cell proliferation / stem cell cycle regulatory responses to exercise.

The other five miRs in the group of nine were differentially expressed at rest from W0 to W12, suggesting linkage to the differential exercise training adaptations separate from acute exercise responses. Of these, miR-935 and miR-12296-5p are previously unknown exercise-responsive miRs, with exosomal miR-935 upregulated in fMQ+/CRF+ and exosomal miR-338-5p downregulated in fMQ+/CRF+. Exosomal miR-935 has been shown to protect against oxidative stress in human cardiac progenitor cells (88). miR-935 also targets NOTCH1, part of the NOTCH pathway shown to play a role in the beneficial effects of exercise on hippocampal neurogenesis and cognitive function in mice (89). miR-338-5p targets NRP1, an angiogenic growth factor that is upregulated in response to exercise (90), suggesting a role for miR-338-5p in exercise-induced angiogenesis. It has also been shown to play a role in skeletal muscle in response to hypoxia (91) and inflammation and oxidative stress (92) in rats.

miR-1296-5p and miR-376a-5p were downregulated in muscle of fMQ+/CRF+ responders in response to training. Both have reported links to exercise, with a downregulation of miR-1296-5p in exosomes reported in response to fatiguing freestyle swimming (36), and an upregulation of miR-376a-5p in plasma reported in response to aerobic exercise training (93). miR-376a-5p plays a role in the response to hypoxia and may promote cartilage repair by promoting proliferation and reducing apoptosis (94). miR-181d-5p, previously reported to be downregulated in plasma of female synchronized swimmers (82), was downregulated in muscle of fMQ+/CRF+ in training and in exosomes in the acute training response. miR-181d-5p targets BCL2 and IRS2 and may thus play a role in apoptosis and insulin resistance. A recent report also implicates miR-181d-5p in reducing hypercholesterolemia by targeting PCSK9 (95). In mice, it reportedly also targets KLF6 to modulate apoptosis and inflammation in kidneys after IR injury (96). NEAT1, a lncRNA shown to modulate the exercise-induced improvements in atherosclerosis (97), inhibits oxidative stress in human vascular endothelial cells by activating the miR-181-d-5p/CDKN3 axis (98).

### Summary

In summary, both TRAD and HITT were highly efficacious combined exercise interventions that similarly improved physiological performance, clinically important health benefits and markers of skeletal muscle health and function despite their differences in intensity and session duration. Unique miR signatures were identified across exercise doses, training status, and early-phase detraining for serum-derived exosomes and muscle, with additional differences noted due to biological sex. We implemented MCIDs to establish individual exercise responsiveness on two key health benefits and employed HOSVD machine learning to map biocircuitry associated with inter-individual response heterogeneity that integrated over 37,000 phenomic and molecular features followed by rigorous down-selection of key features. Cross-validation analysis of DGE and HOSVD yielded a core signature of nine miRs linked to exercise responsiveness. We believe the interrogation undertaken here provides advances in: (i) exercise dose-response physiology; (ii) orchestrated early-phase detraining responses; and (iii) the pursuit of mechanisms to explain inter-individual exercise response heterogeneity that may be leveraged in the future to optimize and individualize exercise prescriptions.

## Data Availability

All data produced in the present study are available upon reasonable request to the authors.

## DATA AVAILABILITY

This trial was registered at ClinicalTrials.gov (NCT03380923). The overall trial design is summarized at ClinicalTrials.gov under the protocol identifier NCT03380923 and is available at https://clinicaltrials.gov/study/NCT03380923?id=NCT03380923%20&rank=1.

Raw small RNA-Seq data for all muscle and exosome miR analysis have been deposited in the Gene Expression Omnibus (GEO) database [GSE269533] and are publicly available as of the date of publication. Any additional information required to reanalyze the data reported in this paper is available from the lead contact upon request.

## SUPPLEMENTAL MATERIAL

Figures S1-S4, Table S1, S3

Tables S2, S4 (Excel files containing additional data too large to fit in a PDF, related to figures 3 and 4).

## ACKNOWLEDGMENTS

We thank all the participants for their hard work and commitment to the trial. We also greatly appreciate the effort of study coordinators, interventionists and biospecimens collection and processing staff.

Figure 5 was created in part using Biorender (license available: https://BioRender.com/a09x082)

## AUTHOR CONTRIBUTIONS

**Conceptualization**, TJB, MMB; **Methodology**, MMB, TJB, IA, JSM, KML, AM; **Software**, JSM, MPC, ZAG, KML, KP, AM; **Validation**, MMB, JSM, MPC, MK, YW; **Formal Analysis**, JSM, MC, GRC, MMB, ZAG, AM; **Investigation**, JSM, MPC, BP, SCT, RSS, KML, ZAG, ABS, SMO, SY, DJD, CJK, KP, MBB, JM, IA, YW, GM, GRC, SP, TJB, MK, MMB; **Resources**, TJB, MMB, MK; **Data Curation**, JSM, MPC, CJK, SY, SMO, KML, BP, YW; **Writing: Original Draft**, JSM, MMB, MPC, ZAG; **Editing and revising**, JSM, MPC, BP, SCT, RSS, KML, ZAG, ABS, SMO, SY, DJD, CJK, KP, MBB, IA, YW, GM, GRC, SP, TJB, MK, MMB; **Final approval**, all authors; **Visualization**, JSM, MPC, MMB, AM; **Supervision**, TJB, MK, MMB, SCT; **Project Administration**, SCT, ABS, TJB, MMB, MK; **Funding Acquisition**, TJB, MMB.

