## Supplemental Tables and Figures for "Multidimensional biocircuitry of exercise adaptation: integrating *in vivo* and *ex vivo* phenomics with miRNA mapping"

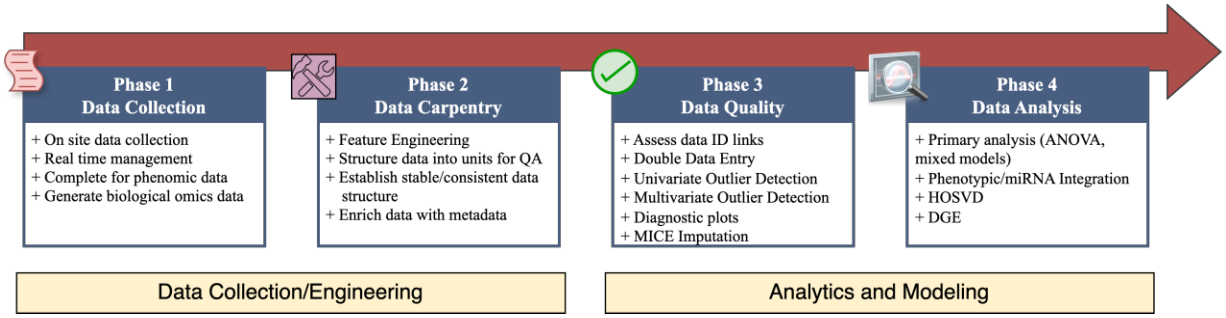


**Supplemental Figure 1. Four phase data management and analysis pipeline.** This is the conceptual pipeline design. The pipeline was implemented in R, using Targets. QA: Quality analysis; ID links: participant and session identifiers; MICE: multiple imputation by chained equations; ANOVA: analysis of variance; Mixed models: mixed effects models; HOSVD: Higher Order Singular Value Decomposition; DGE: Differential Gene Expression.

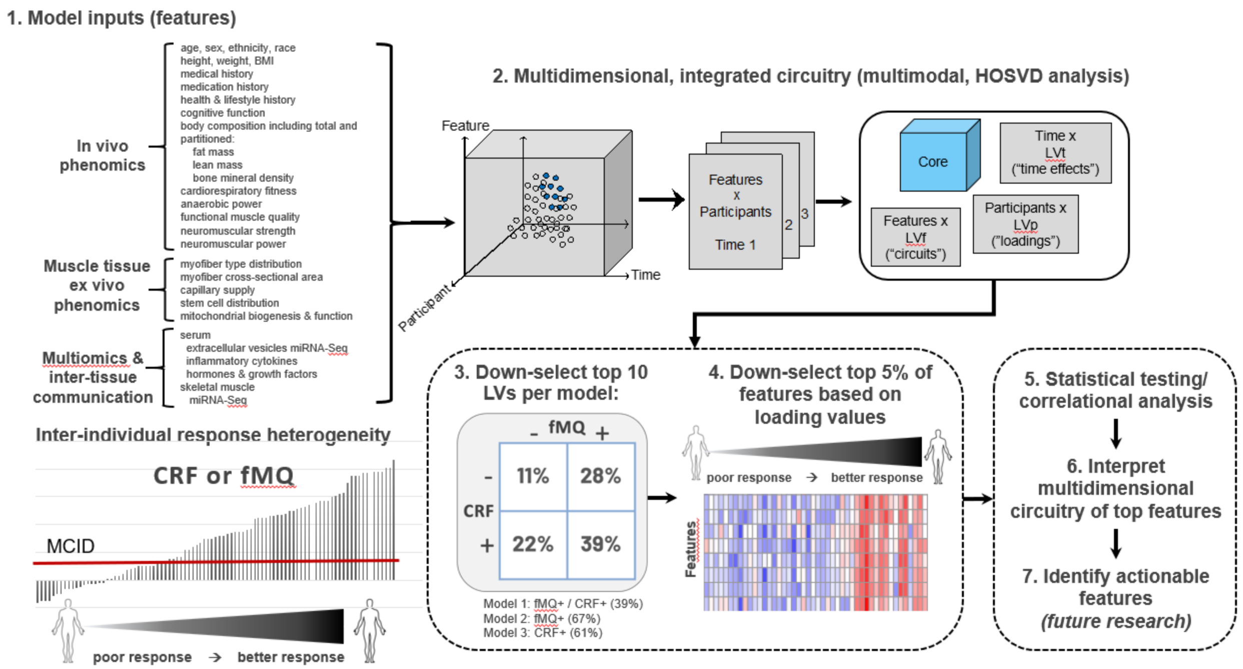


**Supplemental Figure 2: Multidimensional modeling/learning framework.** HOSVD: Higher order singular value decomposition; LV's: latent variables; Punnett square displaying the observed distribution of minimum clinically important difference (MCID) scores for functional muscle quality (fMQ) and cardiorespiratory fitness (CRF); achieved MCID (+); did not achieve MCID (-).

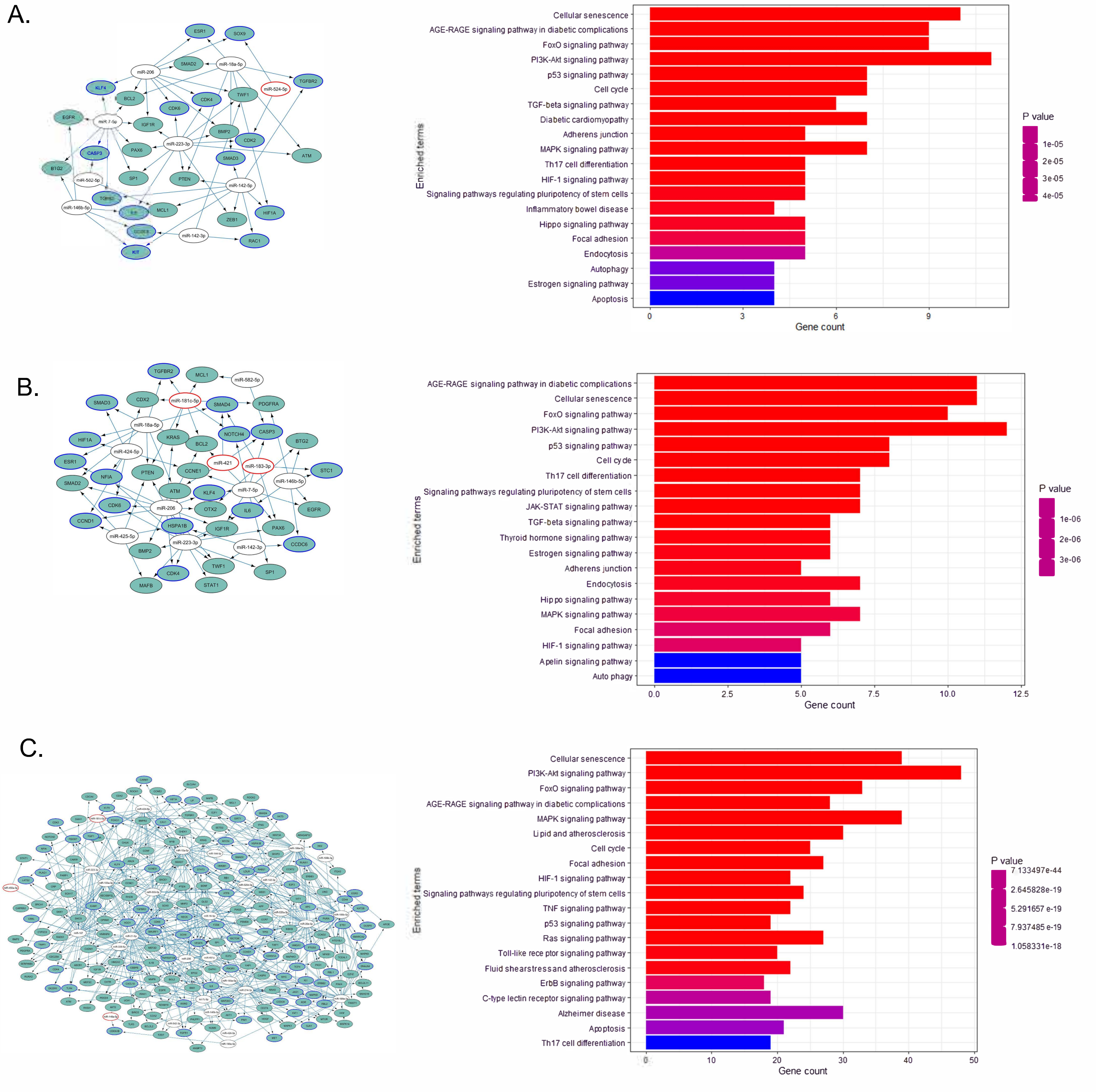


**Supplemental Figure 3: Cytoscape networks and KEGG pathways analysis of DE miRNA of acute exercise responses in muscle for untrained (W0), trained (W12) states and in detraining (W16).** (A-C) Signaling networks (left) generated from DE miRNA differentially expressed in the (A) untrained W0 (W0 3h and W0 24h vs. W0 pre), (B) trained (W12 3h and W12 24h vs. W12 pre), and (C) detrained states (W16 rest vs. W12 pre) showing high-confidence mRNA targets regulated by at least 2 DE miRNA in the dataset. miRNA with red outlines are miRNA with no known link to exercise. mRNA nodes with blue outlines are indicated as exercise-responsive mRNA in MetaMEx. mRNA targets were subjected to KEGG pathway enrichment analysis using Enrichr in R (right). The top 20 enriched non-disease pathways are shown.

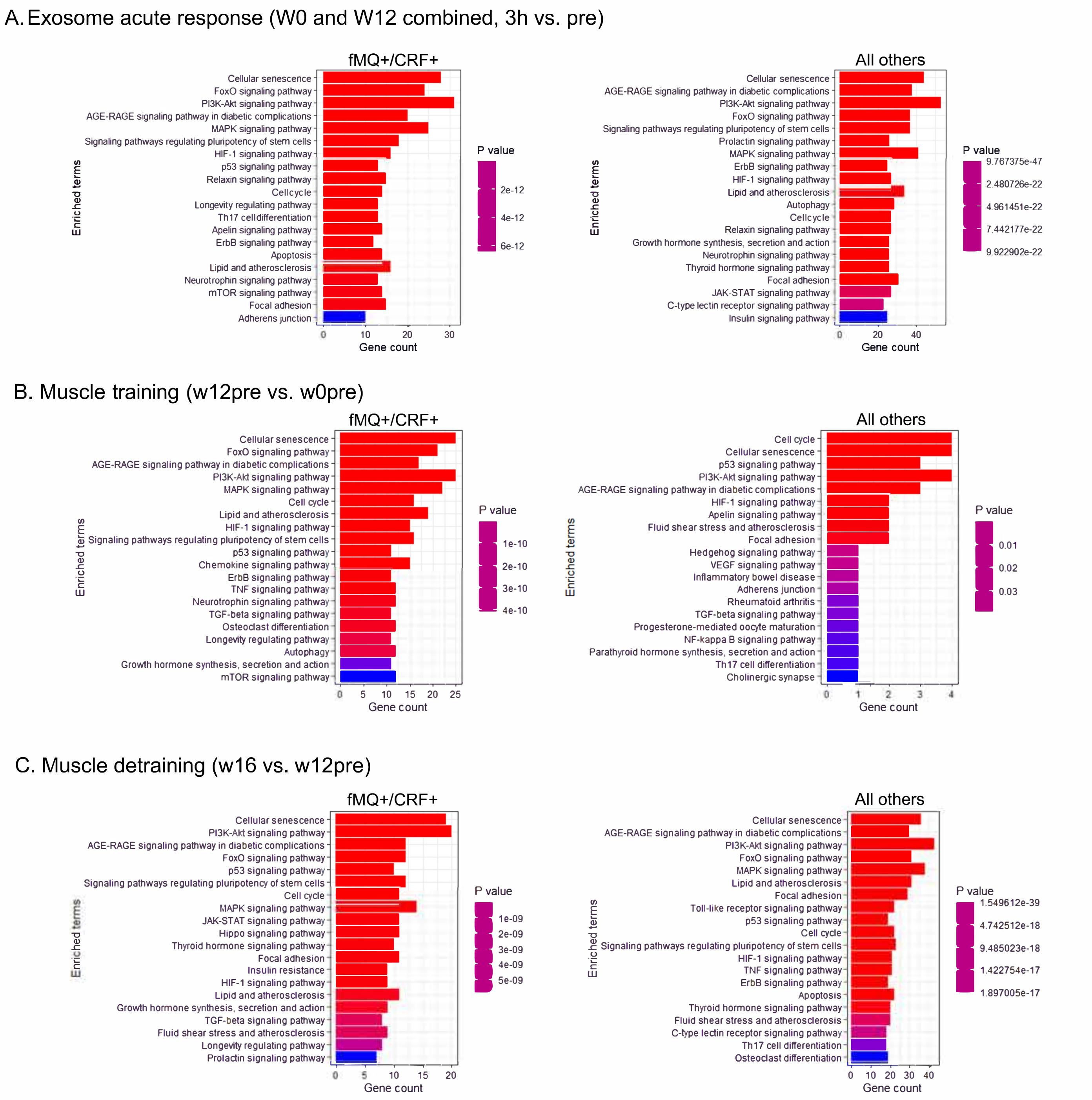


**Supplemental Figure 4: KEGG pathway analysis of high confidence mRNA targets of miRNA DE in CRF+/fMQ+ participants.** High confidence mRNA targets of the DE miRNA from **Figure 3** were identified using miRTarbase v9. These mRNA were subjected to KEGG pathway enrichment analysis using Enrichr in R. The top 20 enriched non-disease pathways are shown for the CRF+/fMQ+ responders (left) and all other participants (right). Pathway enrichment plots for (A) mRNA targes of miRNA DE in the acute exosome response (combined W0 and W12 mRNA target lists of miRNA DE between 3h and pre samples), (B) mRNA targets of miRNA DE in response to training in exosomes (w12pre vs. w0pre), and (C) mRNA targets of miRNA DE in detraining (w16 vs w12pre) are shown.

**Supplemental Table 1.** Resources table. List of key tools and resources used to conduct the study.

| REAGENT or RESOURCE | SOURCE | IDENTIFIER |
|---|---|---|
| **Antibodies** | | |
| Myosin Heavy Chain I (mouse IgG2b) | Developmental Studies Hybridoma Bank | BA-D5 |
| Myosin Heavy Chain IIa (mouse IgG1) | Developmental Studies Hybridoma Bank | A4.74s |
| PAX7+ (mouse IgG) | Developmental Studies Hybridoma Bank | PAX7 |
| Laminin (rat IgG) | ThermoFisher | MA1-06100 |
| Ulex Europaeus Agglutinin I (UEA-1) | Vector Labs | L-1060 |
| AlexaFluor 488 secondary (anti-mouse IgG2b) | ThermoFisher | A-21145 |
| AlexaFluor 488 tyramide SuperBoost Kit, streptavidin | ThermoFisher | B40932 |
| AlexaFluor 594 secondary (anti-mouse IgG1) | ThermoFisher | A-21121 |
| AlexaFluor 647 secondary (anti-rat IgG) | ThermoFisher | A-21247 |
| Biotin-conjugated goat anti-mouse secondary | Jackson ImmunoResearch | 115-065-003 |
| **Chemicals and reagents** | | |
| VectaShield mounting media with DAPI | Vector Labs | H-1200-10 |
| 1x PBS (pH 7.4) | Fisher Scientific | BP243820 |
| Gibco Goat serum | Fisher Scientific | PCN5000 |
| 2.5% Normal Horse Serum | Vector Labs | S201250 |
| **Critical commercial assays** | | |
| MSD V-Plex Pro-Inflammatory Panel 1 | MesoScale Discovery | K15049D |
| Testosterone Tosoh AIA automated immunoassay | TOSOH Bioscience | 025204 |
| Dehydroepiandrosterone (DHEA) Tosoh AIA automated immunoassay | TOSOH Bioscience | 025222 |
| Sex-hormone binding globulin Tosoh AIA automated immunoassay | TOSOH Bioscience | 025239 |
| Androstenedione Tosoh AIA automated immunoassay | TOSOH Bioscience | 025221 |
| Human FGF-21 ELISA | Millipore | EZHFGF21-19K |
| IGF-1 ELISA | ALPCO | 22-IGFHU-E01 |
| IGFBP-3 ELISA | ALPCO | 22-BP3HU-E01 |
| miRVana Paris Kit (muscle total RNA isolation) | ThermoFisher | AM1556 |
| Total exosome isolation and Exosome RNA/Protein Isolation Kit | Life Technologies | 4478545 |
| Agilent 2100 Bioanalyze DNA1000 chip | Agilent | 5067-4626 |
| Ion P1 chip v3 with Ion PI Seq 200 kit v3 | Life Technologies | 4488315 |
| **Deposited data** | | |
| miR fastq/read matrices | Gene Expression Omnibus | GSE269533 |
| **Software and algorithms** | | |

| | | |
|---|---|---|
| Partek Flow v12.0 | Partek | N/A |
| R | R Development Core Team | www.r-project.org |
| R Package: Targets[80] | Targets | https://cran.r-project.org/web/packages/targets/index.html |
| R Package: Tidyverse[81] | Tidyverse | https://cloud.r-project.org/web/packages/tidyverse/index.html |
| R Package: lme4[82] | lme4 | https://cran.r-project.org/web/packages/lme4/index.html |
| R Package: ez[83] | ez | https://cran.r-project.org/web/packages/ez/ez.pdf |
| Limma | Bioconductor | https://bioconductor.org/packages/release/bioc/html/limma.html |
| **Other** | | |
| Ensure Plus liquid protein supplement (8 oz) | Abbott Labs | 64905 |

**Supplemental Table 2: DE miRNA details.** *Due to the length of this table, it is provided as an Excel file (Table_S2.xlsx).* DE miRNA lists used to generate the UPSET plots in Figures 2 and 3 are listed. Each entry lists the tissue type (muscle or serum exosomes), sampling timepoint details (week 0 or 12, hour pre, 0, 3 or details of the comparison (numerator and denominator used), miRNA ID, p value and FC. Also included is a manual curation of any published link to exercise in human studies, the tissues in which the link was reported ("tissue source"), and relevant PMIDs. Reported tissue links are coded as 1 = serum, 2=plasma, 3=muscle, 4=blood, 5=exosome, 6=other.

**Supplemental Table 3. NDEx library information for Cytoscape networks in McAdam et al.** Online interactive versions of the signaling networks shown. Online networks are hosted on NDEx[1] and accessible at the links provided. The interactive networks include DE miRNAs from the indicated figure panel and the corresponding high confidence mRNA targets (*i.e.*, those with experimental supporting evidence from Western blot, microarray or RT-PCR) that are regulated by at least 2 miRNAs. miRNAs with no prior published link to exercise are bordered in red. mRNAs with MetaMEx reported exercise-responsiveness are bordered in blue.

| Network | UUID | NDEX Link | Description |
|---|---|---|---|
| **Figure 4E (left)** Exosome fMQ+CRF+ W0 acute | 183da015-b73b-11ef-99aa-005056ae3c32 | Figure 4E_left | miRNA-mRNA signaling network of the untrained acute response to exercise in fMQ+/CRF+ participants in serum exosomes. miRNA shown are differentially expressed in the acute untrained state (W0, 3h) relative to baseline (week 0 pre). |
| **Figure 4E (right)** Exosome fMQ+CRF+ W12 acute | 72594098-b73b-11ef-99aa-005056ae3c32 | Figure 4E_right | miRNA-mRNA signaling network of the trained acute response to exercise in fMQ+/CRF+ participants in serum exosomes. miRNA shown are differentially expressed in the acute trained state (W12, 3h) relative to baseline (week 12 pre). |
| **Figure 4G** Muscle fMQ+CRF+ Training | d96e699a-b73b-11ef-99aa-005056ae3c32 | Figure 4G | miRNA-mRNA signaling network of the training response to exercise in fMQ+/CRF+ participants in vastus lateralis muscle. miRNA shown are differentially expressed in the trained baseline (W12 pre) relative to untrained baseline (W0 pre). |
| **Figure 4I** Muscle fMQ+CRF+ Detraining | 5bdbd8ed-b73c-11ef-99aa-005056ae3c32 | Figure 4I | miRNA-mRNA signaling network of the detraining response to exercise in fMQ+/CRF+ participants in vastus lateralis muscle. miRNA shown are differentially expressed in the detrained sample (W16) relative to trained baseline (W12 pre). |
| **Figure 6B** 9miRNA common to DGE and HOSVD | 73babb73-bd52-11ef-99aa-005056ae3c32 | Figure 6B | miRNA-mRNA signaling network of the 9 miRNA in the output of the DGE and HOSVD analysis. miRNA shown are differentially expressed in the DGE analysis by MCID and in the HOSVD output. |
| **Figure S3A** Muscle 90-subj untrained | 758bf811-b73d-11ef-99aa-005056ae3c32 | Figure S3A | miRNA-mRNA signaling network of the untrained (W0) response to exercise in the overall exercise response (90 subject dataset). miRNA shown are differentially expressed in the untrained state (W0 3h vs W0 pre and W0 24h vs W0 pre). |
| **Figure S3B** Muscle 90-subj trained | a5759813-b73d-11ef-99aa-005056ae3c32 | Figure S3B | miRNA-mRNA signaling network of the trained (W12) response to exercise in the overall exercise response (90 subject dataset). miRNA shown are differentially expressed in the trained state (W12 3h vs W12 pre and W12 24h vs W12 pre). |
| **Figure S3C** Muscle 90-subj detrained | f351d855-b73d-11ef-99aa-005056ae3c32 | Figure S3C | miRNA-mRNA signaling network of the detrained (W16) response to exercise in the overall exercise response (90 subject dataset). miRNA shown are differentially expressed in the detrained state (W16) relative to the trained baseline (W12 pre). |

**Supplemental Table 4: ENRICHR pathway analysis table for McAdam *et.al.*** *Due to the length of this table, it is provided as an Excel file (Table_S4.xlsx).* The full list of KEGG 2021 non-disease enriched pathways ($p<0.05$) resulting from ENRICHR analysis for Figures 4E, 4G, 5C, S2 and S3 are provided. For each, the enriched pathway, overlap, p-value, adjusted p-value, odds ratio, combined score and genes appearing in the indicated pathways are provided. mRNA lists used for ENRICHR analysis are provided in the “mRNA target lists” tab.
